# Unraveling COVID-19: Descriptive Analytics in a Middle-Income Country, Paving the Path Forward

**DOI:** 10.1101/2023.08.16.23294160

**Authors:** Norvin P. Bansilan, Jomar F. Rabajante

## Abstract

The outbreak of COVID-19 unleashed an unprecedented global pandemic, leaving a profound impact on lives and economies worldwide. Recognizing its severity, the World Health Organization swiftly declared it a public health emergency of international concern. Tragically, the Philippines reported the first death case outside China, leading to a surge in cases following the first instance of local transmission. In response to this crisis, collaborative efforts have been underway to control the disease and minimize its health and socio-economic impacts. The COVID-19 epidemic curve holds vital insights into the history of exposure, transmission, testing, tracing, social distancing measures, community lockdowns, quarantine, isolation, and treatment, offering a comprehensive perspective on the nation’s response. One approach to gaining crucial insights is through meticulous analysis of available datasets, empowering us to inform future strategies and responses effectively. This paper aims to provide descriptive data analytics of the COVID-19 pandemic in the Philippines, summarizing the country’s fight by visualizing epidemiological and mobility datasets, revisiting scientific papers and news articles, and creating a timeline of the key issues faced during the pandemic. By leveraging these multifaceted analyses, policymakers and health authorities can make informed decisions to enhance preparedness, expand inter-agency cooperation, and combat future public health crises effectively. This study seeks to serve as a valuable resource, guiding nations worldwide in comprehending and responding to the challenges posed by COVID-19 and beyond.

## 1. Introduction

In early December 2019, an outbreak of severe acute respiratory syndrome coronavirus-2 (SARS-CoV-2) was reported in Wuhan, China. Initially thought to be a zoonosis, it was later recognized that the significant spread of SARS-CoV-2 is primarily attributed to human-to-human transmission [1]. Following exposure to the virus, individuals may experience symptoms such as fever, cough, sore throat, loss of taste or smell, difficulty breathing, and diarrhea, with an incubation period of 2 to 14 days [2].

The outbreak quickly spread to various countries, including Canada, France, Germany, India, Italy, Japan, Singapore, South Africa, South Korea, Spain, the United Kingdom, the United States, Vietnam, and the Philippines. Consequently, the World Health Organization (WHO) declared a global public health emergency, officially naming the novel coronavirus (nCoV) disease ‘COVID-19’, short for COronaVirus Disease-2019 [3] [4] [5].

In response, the Philippines, like many other nations, closely monitored the outbreak’s development and implemented measures to curb its spread. However, despite their efforts, the country reported its first confirmed death outside of China from COVID-19 on February 2, 2020, marking a solemn moment for the nation and the global community [6]. Subsequently, intensified containment efforts were undertaken, involving travel restrictions, quarantine measures, and public health campaigns. The surge in cases following the first local transmission necessitated enhanced contact tracing, widespread testing, and strict community lockdowns [7].

The Philippine government responded by increasing hospital beds and healthcare services, albeit at the expense of other diseases. Additionally, the government procured vaccines under Emergency Use Authorization through international collaborations, with mass vaccination campaigns targeting healthcare workers and vulnerable groups to pave the way for a return to normalcy [8] [9]. The collaborative efforts of the government, health workers, researchers, data analysts, citizens, and various sectors yielded optimistic results in combating the spread of the disease.

In May 2023, the WHO declared COVID-19 no longer a global public health emergency [10]. To facilitate the conclusion of the pandemic and aid the country in moving forward, this paper provides descriptive data analytics on COVID-19 in the Philippines. Intending to serve as a concise reference for future research, the study offers summaries and visualizations of epidemiological and related datasets. The research also includes a collection of scientific papers and news articles that studied and reported on the disease’s status in the country, along with a timeline of key events throughout the pandemic. Through these comprehensive analyses, this paper seeks to contribute valuable insights to the broader scientific community, guiding future pandemic responses and preparedness efforts.

## 2. COVID-19 Data Dashboard

In this paper, we introduce the Philippine COVID-19 dashboard, developed using Google LookerStudio [11]. The datasets utilized for the dashboard were predominantly gathered from the official Department of Health (DOH), Philippines website as of 28 April 2023 [12]. The dashboard offers a comprehensive range of insights, including national, regional, and provincial epidemic curves and mortality trends, testing dynamics, age and sex profiles, severe and critical cases, and cases in the Intensive Care Unit (ICU). Additionally, the dashboard presents data on vaccination progress, the global epidemic curve, and Philippine Google mobility trends [13] [14] [15].

Each section within the dashboard incorporates data visualization tailored to its specific profile, providing an interactive and informative experience. As a preview, this paper showcases selected figures from the dashboard, offering a glimpse of the extensive data visualization and analytical capabilities it provides.

Figure 1 illustrates the timeline of key COVID-19 events, along with the national epidemic curve (depicted in blue) and cumulative count of COVID-19 cases (depicted in cyan), from January 2020 to April 2023. The months are abbreviated as J, F, M, A, M, J, J, A, S, O, N, and D. Additionally, the green arrows indicate the country’s mo-bility trends, with a downward trend indicating a decrease in average mobility and an upward trend suggesting an increase in average mobility. However, note that the upward trend is not monotonic, as there were events where the government imposed mobility restrictions for specific periods, depending on the epidemiological situation.

**Figure 1:**
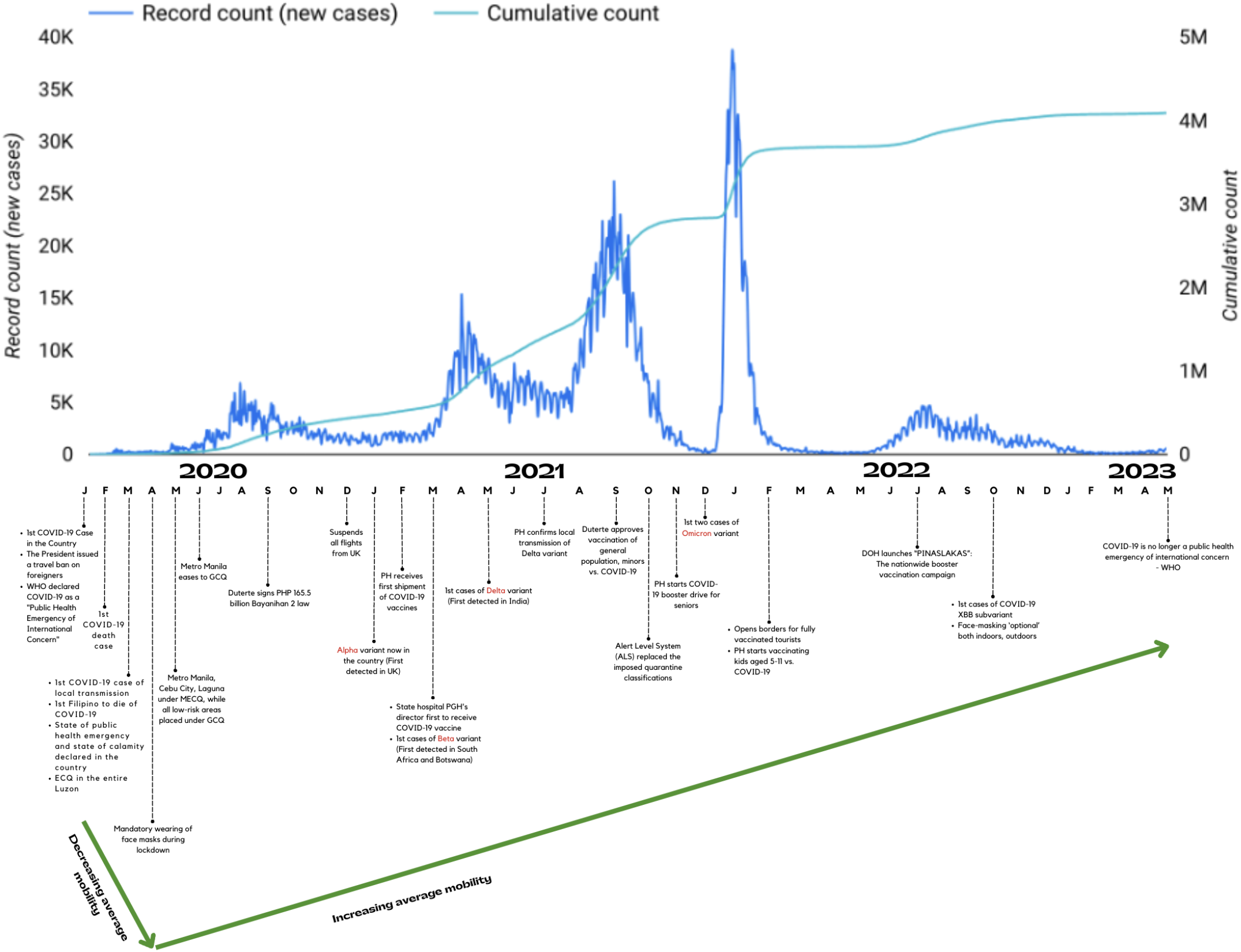
Timeline of Significant Events in the Philippines’ National COVID-19 Epidemic Curve. The x-axis represents the initial of the months from 2020 to 2023, while the left and right y-axes denote the COVID-19 record count of new cases and cumulative count of cases, respectively. The case count is based on the confirmation or reporting date and is not corrected to consider delays in the testing process and the presence of undetected cases.

During the initial months of January to April 2020, Figure 1 displays a decline in average mobility and a low number of COVID-19 cases. These changes were likely influenced by the government’s imposition of lockdown measures to curb the spread of the virus [16] [17]. Subsequently, starting from April 2020 up to the present, the figure demonstrates an increase in average mobility, coinciding with a rise in COVID-19 cases. Notably, July and August 2020 witnessed a peak of 6,836 daily infections on August 10, 2020. This surge can be attributed to the easing of quarantine and restrictions in May and June 2020 [18] [19]. Further spikes in cases occurred in March and April 2021, reaching a peak of 15,252 daily infections on April 2, 2021. These increases were likely influenced by the presence of the Alpha and Beta variants in the country, as well as the gradual reopening of schools, increased inbound travel, and the resumption of recreational activities in certain regions [20] [21] [22] [23]. In May and June 2021, there was a slight rise in cases, possibly due to the reported presence of multiple variants (Alpha, Beta, Delta) in the country [24]. Another notable surge occurred in August and September 2021, with a peak of 26,128 daily cases recorded on September 11, 2021. This increase can be attributed to a surge in Delta variant cases and the easing of community quarantine restrictions in the National Capital Region (NCR) and other areas [25] [26]. Following the surge, a gradual decline in COVID-19 cases was observed, likely attributable to the availability and administration of COVID-19 vaccines to both the general population and minors in the country [27]. The highest single-day tally of COVID-19 cases occurred on January 15, 2022, with 38,776 reported cases. This peak is linked to the surge of the COVID-19 Omicron variant in the country [28]. As of the current date, there has been no significant increase in COVID-19 cases recorded after the January 2022 peak. For a more comprehensive timeline and detailed illustration of the country’s mobility trend, please refer to Section 4 and Figure 14, respectively.

Figures 2, 3, and 4 present the epidemic curve of COVID-19 on a global scale, within Western Pacific countries, and among ASEAN countries, respectively. The data reveal significant disparities in the timing and magnitude of COVID-19 case peaks across these regions. The worldwide epidemic curve in Figure 2 shows that the highest number of COVID-19 cases occurred on December 23, 2022, reaching a staggering 7,945,883 confirmed infections. On the same date, the Western Pacific countries, as illustrated in Figure 3, also experienced their peak with 7,210,956 cases, reflecting the severity of the pandemic’s impact in that region. In contrast, the ASEAN countries, depicted in Figure 4, had their highest number of cases recorded on March 12, 2022, reaching a total of 536,690 infected individuals. This distinct peak date highlights the variation in COVID-19 transmission dynamics within the ASEAN region. An important observation from these figures is that the surge of COVID-19 cases in the global, Western Pacific, and ASEAN epidemic curves did not align temporally, indicating that different factors and responses influenced the spread of the virus in each region. The variations in the peaks emphasize the complex and dynamic nature of the pandemic and its impact on different geographical areas.

**Figure 2:**
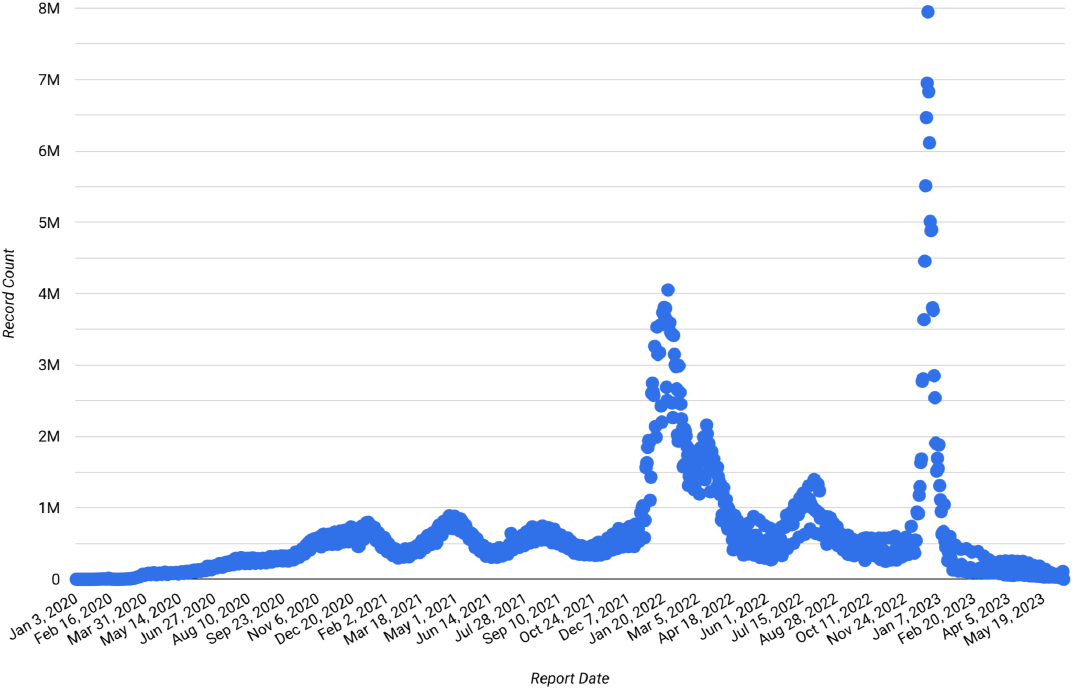
Global COVID-19 Epidemic Curve. The x-axis represents the report date from 2020 to 2023, while the y-axis denotes the number of daily COVID-19 cases.

**Figure 3:**
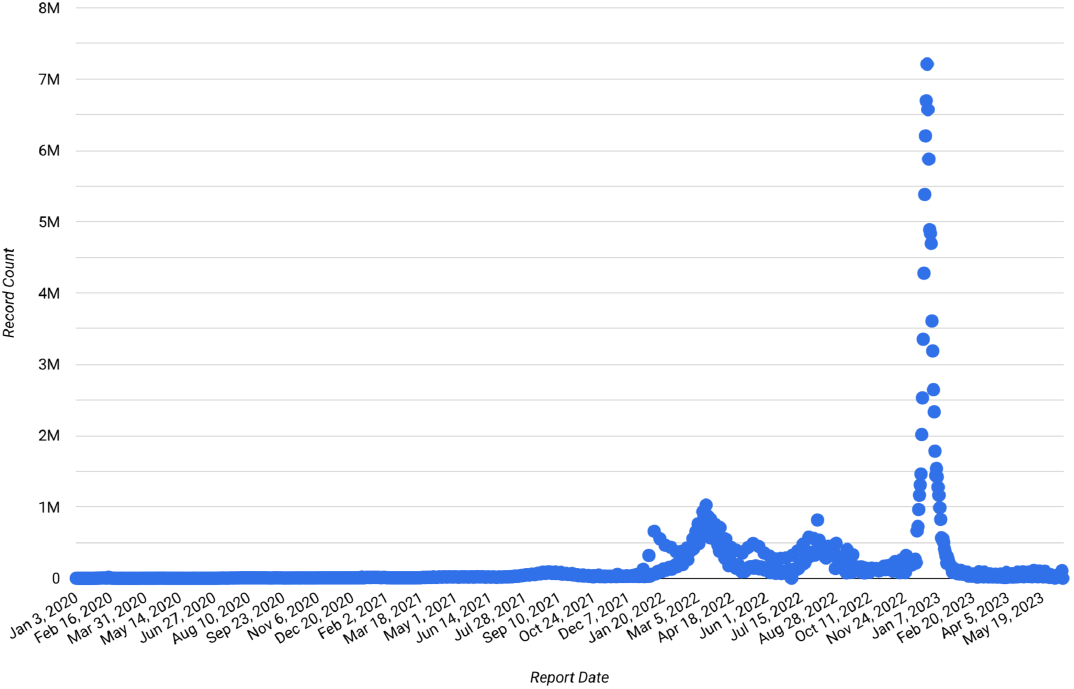
COVID-19 Epidemic Curve in WHO Western Pacific Countries. The x-axis represents the report date from 2020 to 2023, while the y-axis denotes the number of daily COVID-19 cases.

**Figure 4:**
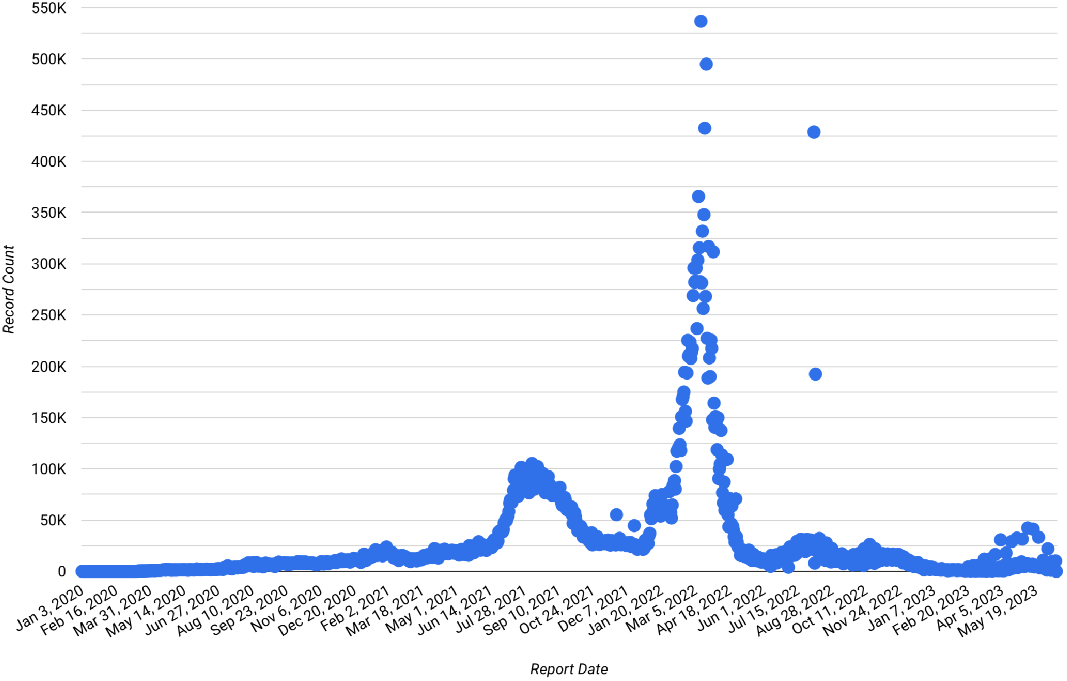
COVID-19 Epidemic Curve in ASEAN Countries. The x-axis represents the report date from 2020 to 2023, while the y-axis denotes the number of daily COVID-19 cases.

Figures 5,6,7, and 8 provide a descriptive detailed analysis of the COVID-19 epidemic curve in some of the most populous regions in the country, namely National Capital Region (NCR), Region 4A (CALABARZON), Region 7 (Central Visayas), and Region 11 (Davao Region), from January 2020 to April 2023. The top figures are with references marked at 10k and 20k daily cases while the bottom figures provide a closer look at specific regions. In line with the national epidemic curve, these regional graphs show that the highest recorded peak of COVID-19 cases across NCR, Region 4A, Region 7, and Region 11 occurred in January 2022. During this time, NCR reported a single-day tally of 18,705 cases, Region 4A documented 9,476 cases, Region 7 recorded 2,514 cases, and Region 11 registered 2,576 cases. This simultaneous peak in multiple large regions indicates synchronicity with the national trend. Furthermore, the figures illustrate that the second-highest peak of COVID-19 cases in these regions was observed during August and September 2021. This similarity in surge timing further supports the semi-synchronous nature of the epidemic curve across the country’s populous regions. However, zooming in there are some asynchronous waves when we compare epidemic curves of the provinces, especially in comparison with the cities (refer to the Google LookerStudio dashboard [11]). Following these peaks, a noteworthy trend emerges where the number of COVID-19 cases started to decrease after reaching the highest levels. Moreover, as of the present date, there has been no high-risk increase in COVID-19 cases reported in these regions.

**Figure 5:**
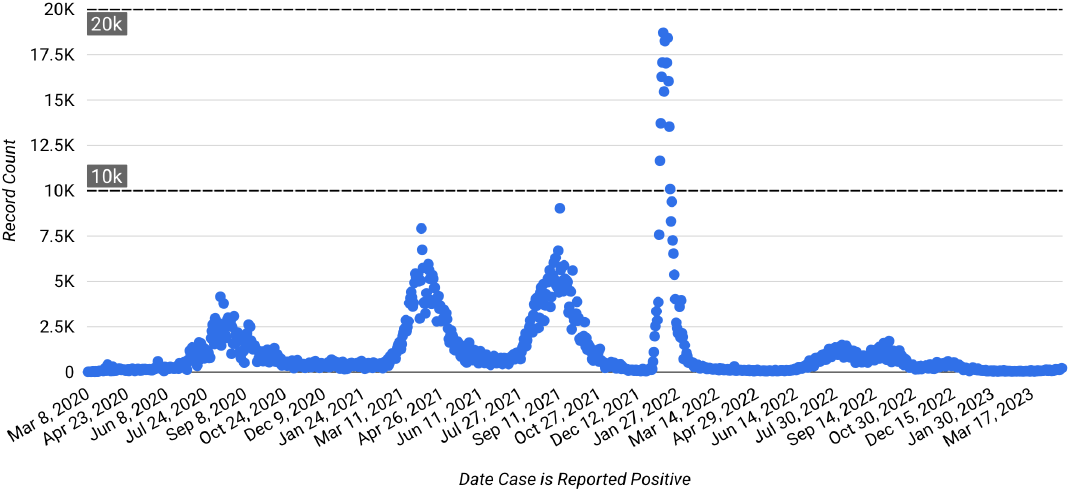
COVID-19 Epidemic Curve in the National Capital Region, Philippines. The x-axis represents the report date from 2020 to 2023, while the y-axis denotes the number of daily COVID-19 cases. References are marked at 10k and 20k daily cases in the top figure.

**Figure 6:**
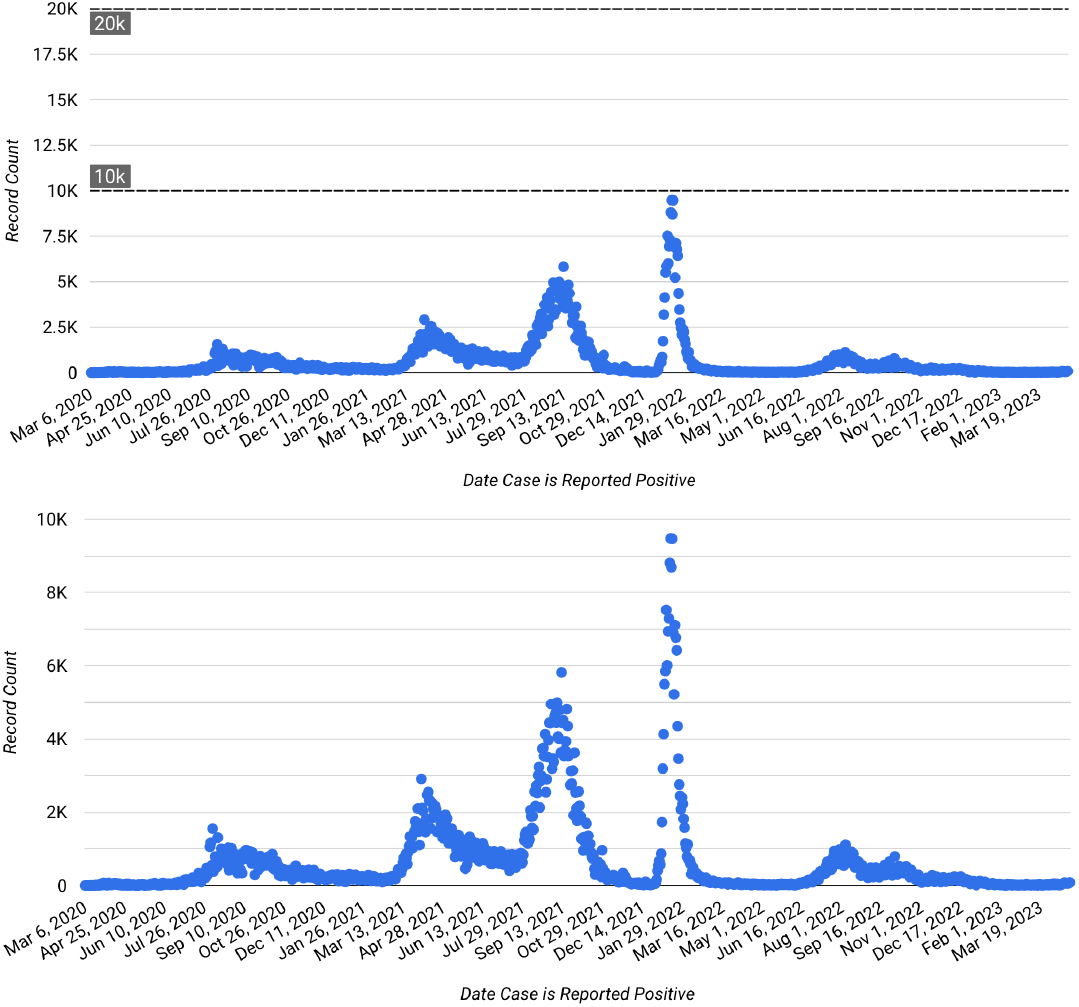
COVID-19 Epidemic Curve in the Region 4A: CALABARZON, Philippines. The x-axis represents the report date from 2020 to 2023, while the y-axis denotes the number of daily COVID-19 cases. References are marked at 10k and 20k daily cases in the top figure.

**Figure 7:**
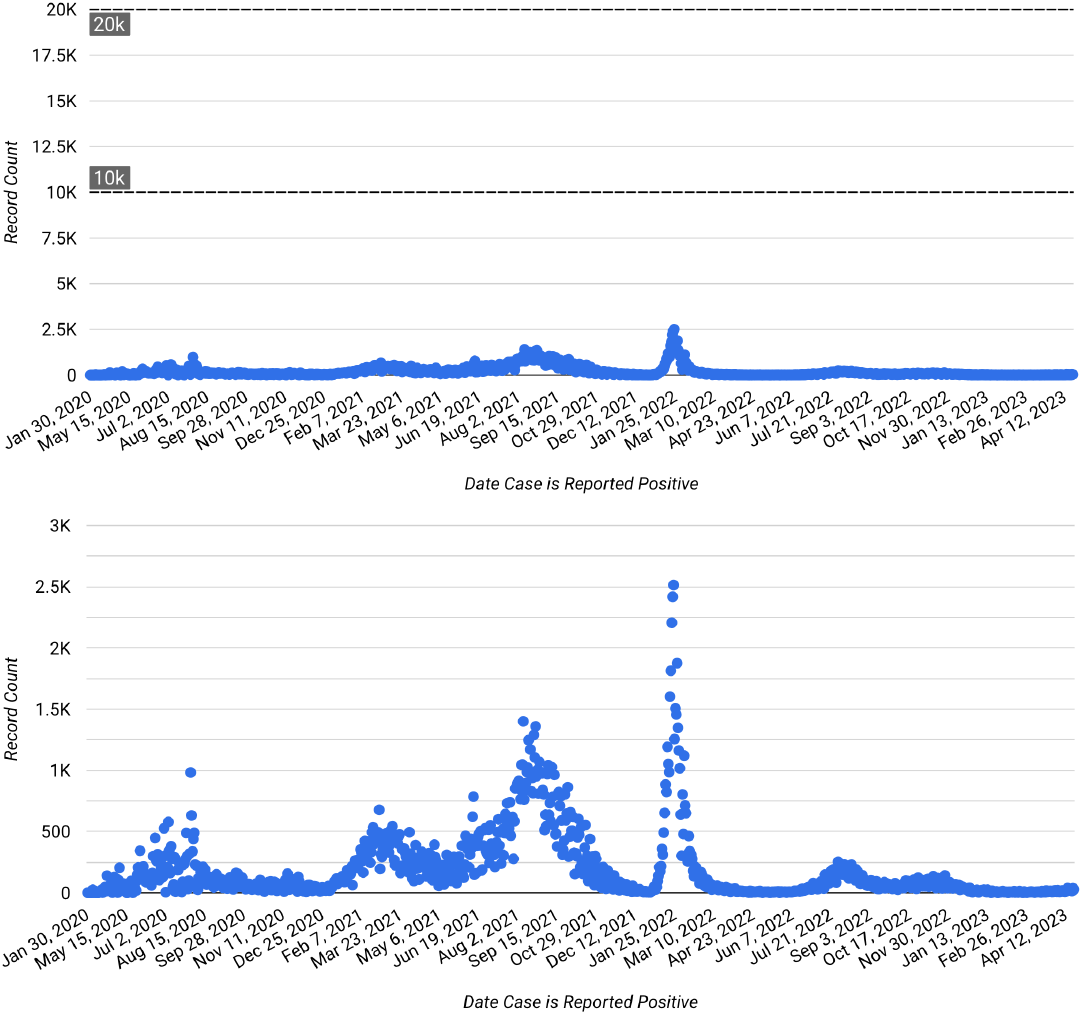
COVID-19 Epidemic Curve in the Region 7: Central Visayas, Philippines. The x-axis represents the report date from 2020 to 2023, while the y-axis denotes the number of daily COVID-19 cases. References are marked at 10k and 20k daily cases in the top figure.

**Figure 8:**
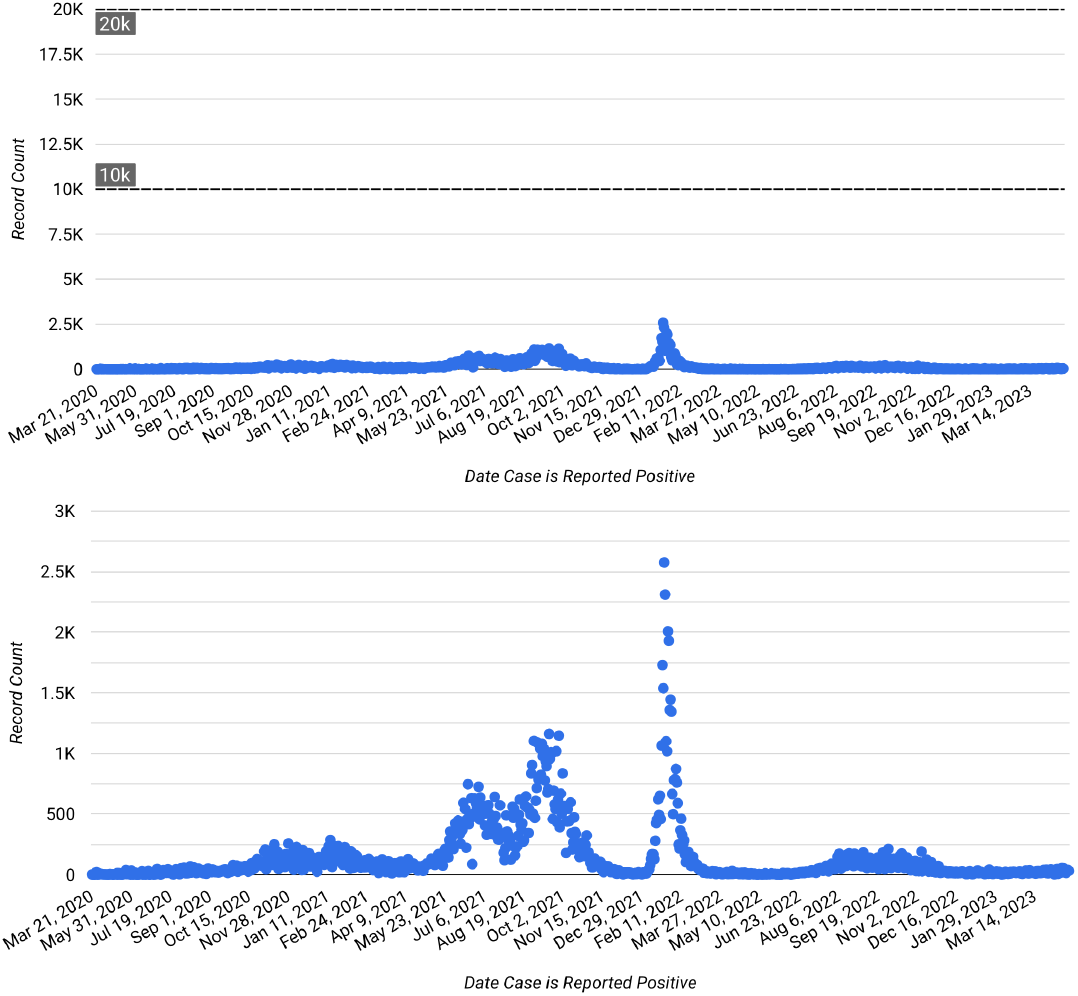
COVID-19 Epidemic Curve in the Region 11: Davao Region, Philippines. The x-axis represents the report date from 2020 to 2023, while the y-axis denotes the number of daily COVID-19 cases. References are marked at 10k and 20k daily cases in the top figure.

The status of COVID-19 patients can range from asymptomatic to critical. Figure 9 presents an insightful view of the varying severity of illness experienced by patients with COVID-19. The graph showcases the number of confirmed severe cases (depicted in blue) and critical cases (shown in red) recorded from 2020 to 2023. Severe cases encompass patients with pneumonia and hypoxemia, while critical cases involve individuals with respiratory failure, septic shock, or multiple organ dysfunction, as stated in the study by Yuki et al. [1]. As highlighted in the figure, there is an occurrence of waves with a high number of severe cases. Its peak occurred on September 10, 2021, with 4,821 confirmed cases. In comparison, critical illness recorded its highest peak on September 12, 2021, with 1,597 confirmed cases. Notably, following these peaks, there has been no high-risk increase in the number of confirmed severe and critical cases. This observation suggests that the biological conditions and the medical efforts to manage and mitigate the impact of the pandemic have been effective in controlling the severity of illness experienced by COVID-19 patients.

**Figure 9:**
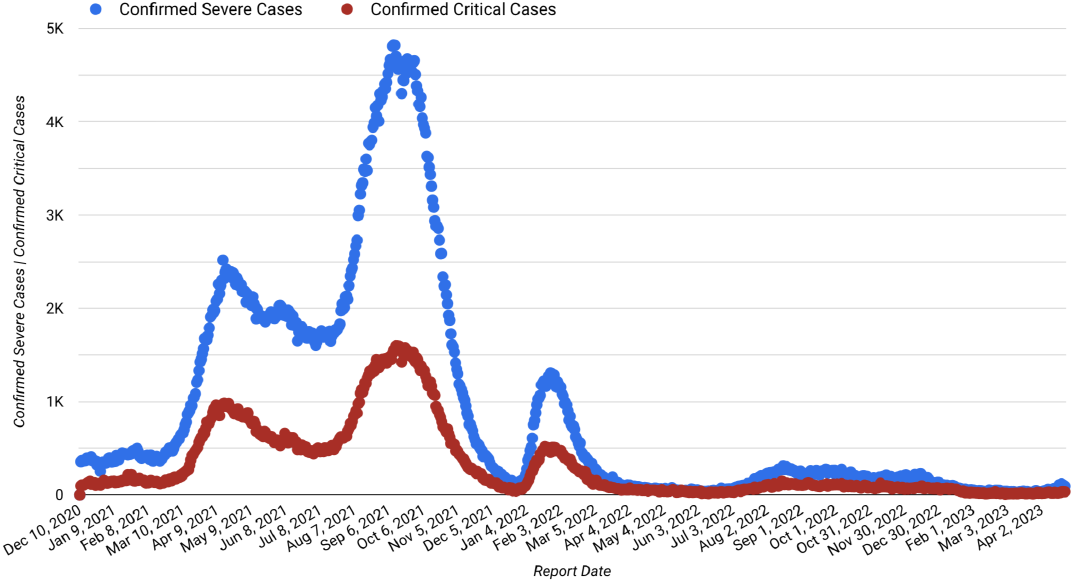
Confirmed Severe Cases and Confirmed Critical Cases in the Philippines. The x-axis represents the report date from 2020 to 2023, while the y-axis denotes the number of confirmed severe and critical cases.

Presenting Figure 10, which provides an overview of the total number of occupied ICU beds in the Philippines from 2020 to 2023, integrating the ICU bed counts per region (top figure). The graph depicts the trends in occupied ICU beds over time, highlighting the variations across different regions. As indicated in the figure, the number of occupied ICU beds increased steadily from 2020 until September 2021. The highest number of occupied ICU beds, totaling 3,438, was recorded on September 22, 2021 (during the “Delta wave”), with the NCR having the most occupied ICU beds (1,272) among all the regions. Following NCR, Central Luzon recorded 415 occupied ICU beds, and CALABARZON had 227 occupied ICU beds on the same day. Conversely, ARMM, MIMAROPA, and SOCCSKSARGEN reported the least number of occupied ICU beds on that date. However, it is encouraging to observe that the number of occupied ICU beds has decreased since September 2021 and continues to be below the 30% occupancy rate up to the present date. This decline could indicate successful efforts in managing COVID-19 cases and the strain on healthcare facilities.

**Figure 10:**
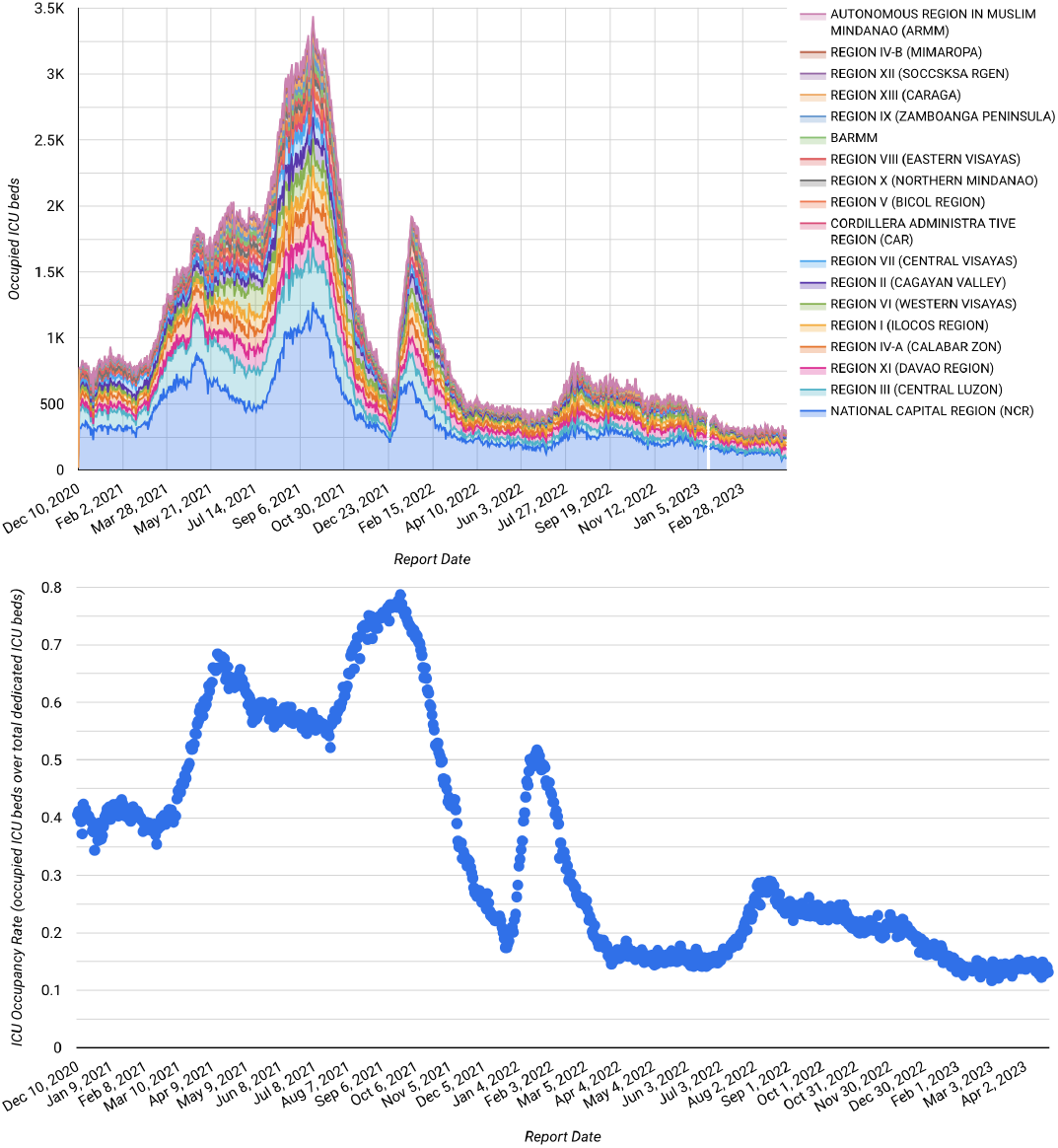
Occupied ICU Beds and ICU Occupancy Rate in the Philippines. The x-axis represents the report date from 2020 to 2023, while the y-axis in the top and bottom figures denote the number of occupied ICU beds and ICU occupancy rate, respectively.

The bottom figure provides a visualization of the ICU bed occupancy rate, calculated by dividing the number of occupied ICU beds by the total dedicated ICU beds. The graph illustrates the fluctuations in the occupancy rate over time. Notably, the highest occupancy rate, reaching 79%, was recorded on September 22, 2021. This spike in the occupancy rate coincided with a significant increase in the number of occupied ICU beds, as demonstrated in the top figure.

Figure 11 depicts the number of COVID-19 death cases in the Philippines recorded throughout the pandemic. Notably, the highest single-day death toll occurred last September 2021, with a tragic count of 442 individuals succumbing to the virus. Remarkably, this peak in fatalities coincided with the highest number of severe and critical patients, as evident in the data. The convergence of these peaks in severe cases and COVID-19-related deaths underscores the significant impact of the pandemic during that period.

**Figure 11:**
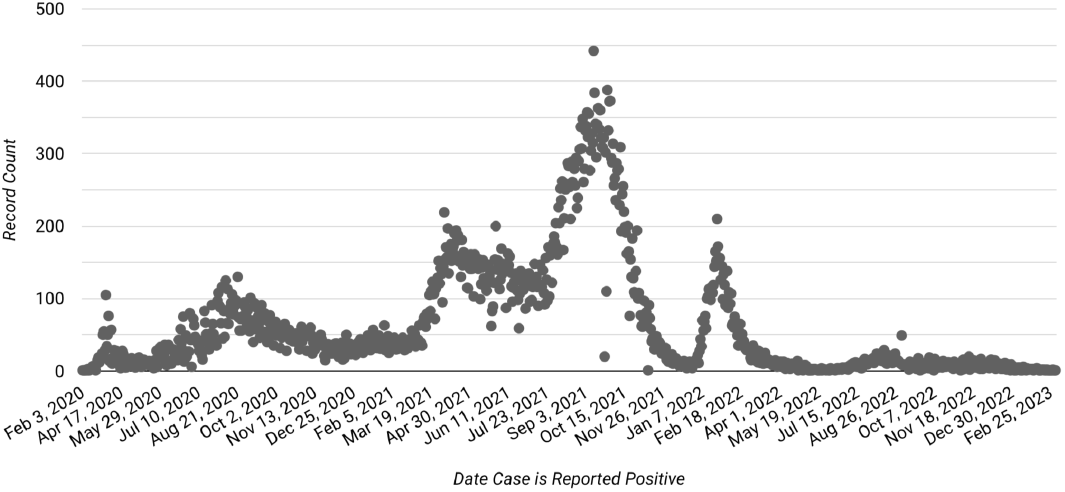
Philippine Death Cases. The x-axis represents the report date from 2020 to 2023, while the y-axis denotes the record count of COVID-19 deaths.

The dynamics of the COVID-19 infected new cases and death cases have started to be decoupled after COVID-19 vaccines have been rolled out and after a new COVID-19 variant which is fast to spread but less likely to cause severe conditions has started to spread. This is evident in the relatively low death cases during the high Omicron wave in January 2022.

In Figure 12, the percentage of deceased individuals (dark gray) and recovered patients (pink) is depicted. The data shows that over 98% of all COVID-19-positive individuals have successfully recovered from the disease, while approximately 1.6% lost their lives due to the illness.

**Figure 12:**
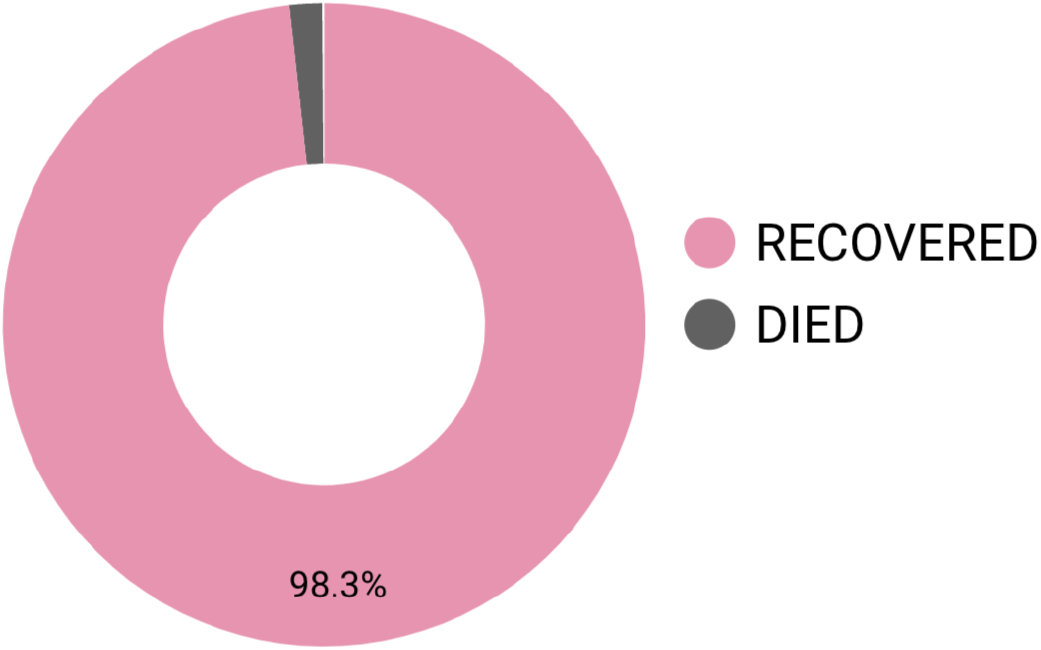
Recovered vs. Died Cases in the Philippines.

Looking at the global deaths, Figure 13 presents the daily count of new COVID-19 deaths (dark gray) and the cumulative deaths (violet). Notably, the period from De-cember 2020 to April 2021 recorded a significant surge in fatalities, with an average of approximately 15,000 to 17,000 death cases per day. Furthermore, an increase in reported deaths was observed in January and February 2022, with around 12,000 to 14,000 death cases per day. The cumulative death toll has reached approximately 7 million from January 2020 to June 4, 2023, illustrating the devastating impact of the pandemic. Notably, there was an evident exponential growth of cumulative deaths from December 2020 to December 2021, highlighting the severity of the crisis during that period. Comparing the global and Philippine situations, there are differences in the dynamics of COVID-19 death counts, especially when the peaks occurred.

**Figure 13:**
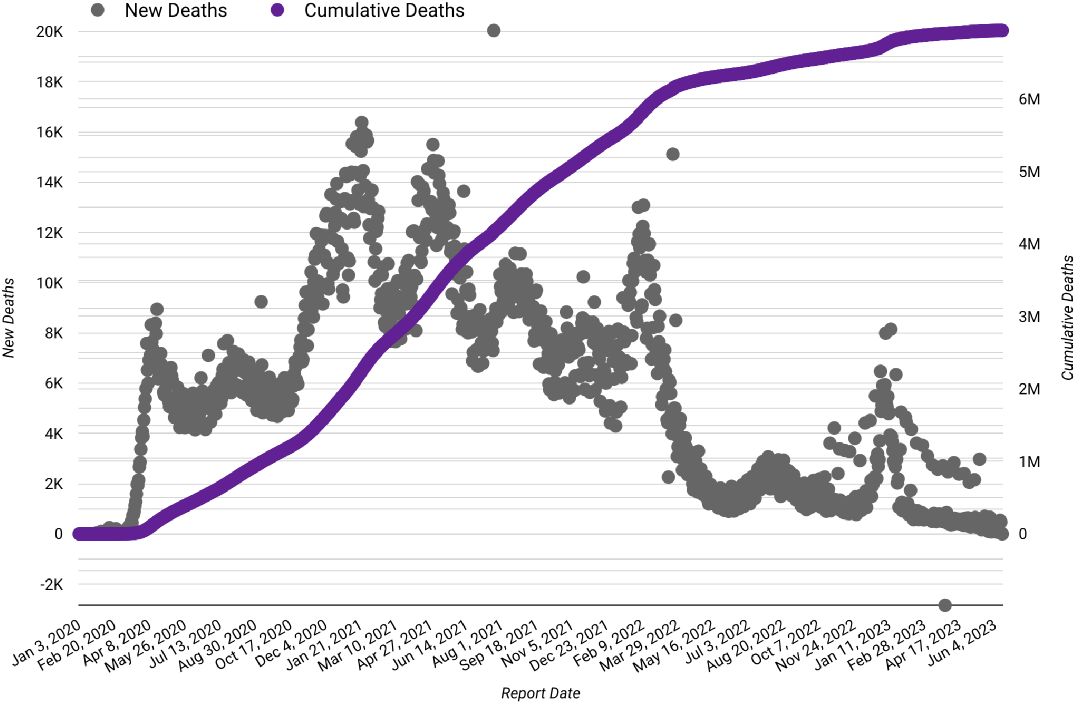
Global New Deaths and Cumulative Deaths. The x-axis represents the report date from 2020 to 2023, while the left and right y-axes denote the record count of COVID-19 new deaths and cumulative deaths, respectively.

Figure 14 illustrates the mobility trends in the Philippines from February 2020 to October 2022. During the initial period from February to April 2020, the mobility trends for the workplace (dark green), parks (yellow), transit stations (orange), retail and recreation (green), and grocery and pharmacy (blue) experienced an exponential decay, with changes ranging from -60% to -80% relative to the baseline. This reduction in mobility can be attributed to the government-imposed lockdown measures. However, starting in May 2020, a gradual increase in mobility was observed across these locations, continuing until October 2022. Notably, the mobility in grocery and pharmacy showed the fastest percentage change from the baseline during this period. Conversely, the mobility trend for residential areas (violet) remained relatively stable, with no significant changes observed except for an initial increase of 30% from the baseline during the first implementation of the lockdown, which was expected as people stayed at home to comply with restrictions.

**Figure 14:**
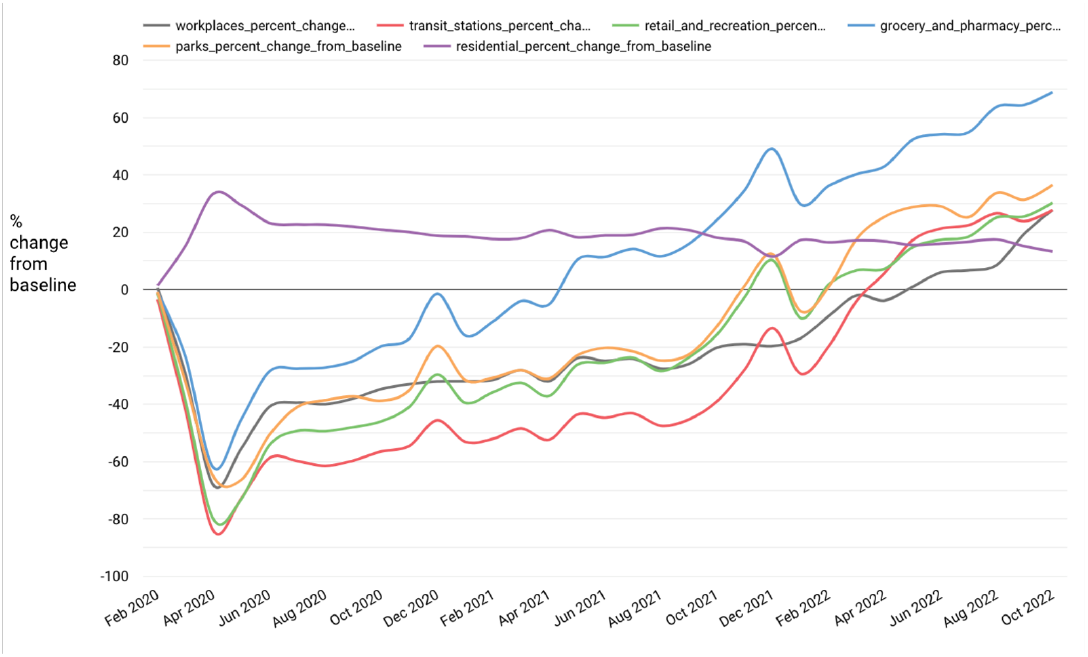
Google Philippine Mobility. The x-axis represents the report date from February 2020 to October 2022, while the y-axis denotes the percentage change from the baseline.

Figure 15 presents the distribution of COVID-19-positive cases across different age groups. The age range of 20 to 40 years exhibits the highest number of infections, with cases ranging from 400,000 to approximately 600,000, notably peaking in the age group from 25 to 29. Conversely, the age groups of 0 to 14 and 65 and above show a lower number of infected cases, ranging from approximately 50,000 to 100,000. These findings highlight the varying exposure and vulnerability of different age groups to the virus, especially relating to the actual age population size and mobility related to age groups.

**Figure 15:**
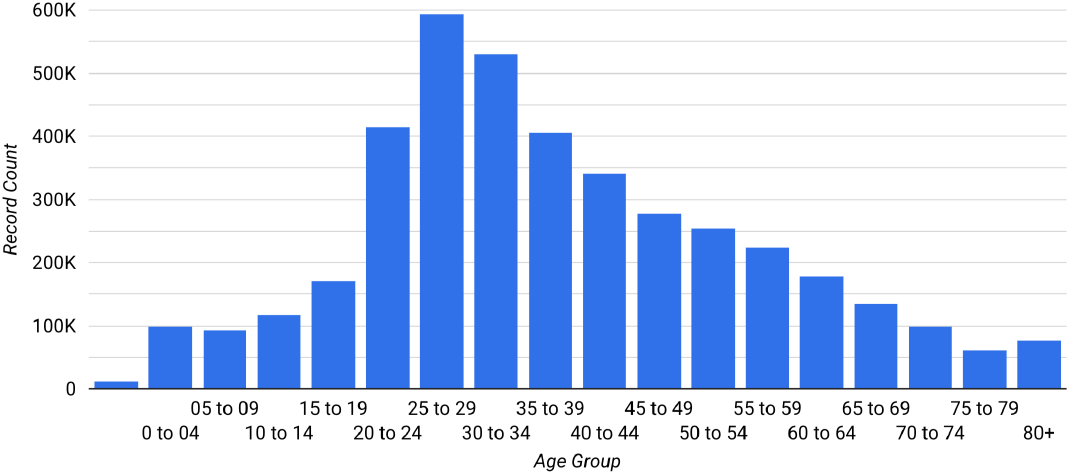
COVID-19 Positive Cases per Age Group in the Philippines. The x-axis represents the age group, while the y-axis denotes the record count of COVID-19-positive cases.

Presented in Figure 16 is the distribution of COVID-19-positive cases based on sex. 51.5% of the cases were females (yellow), slightly outnumbering the 48.5% of cases among males (blue). The disparity in the percentage of infected cases between females and males is marginal.

**Figure 16:**
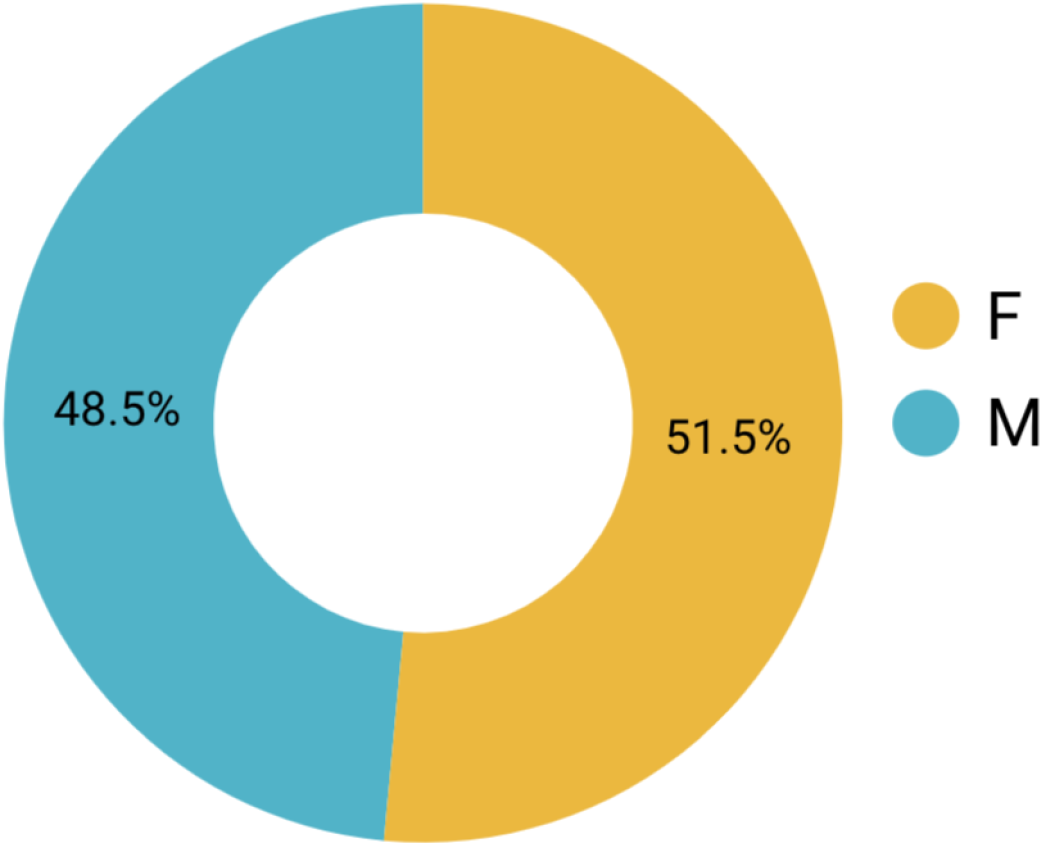
COVID-19 Positive Cases per Sex in the Philippines. Male (M) and Female (F) are depicted in blue and yellow, respectively.

Figure 17 depicts the distribution of COVID-19 cases across distinct age groups, stratified by sex. Notably, the age ranges of 0 to 14 and 35 to 49 exhibit the highest percentage of infected males (blue), with a significant proportion of 53.65% of cases attributed to the age group 0 to 4. Conversely, the age groups of 15 to 34 and 50 and above show the highest prevalence of infected females (yellow), with a substantial 62.36% of cases coming from the 80 and above age group. These observations shed light on the sex-specific variations in infection patterns within different age segments. The difference in the male-female number of cases, especially looking at senior citizens, may be due to the imbalance of actual sex proportions in older groups (e.g., what age group has a longer life span) [29].

**Figure 17:**
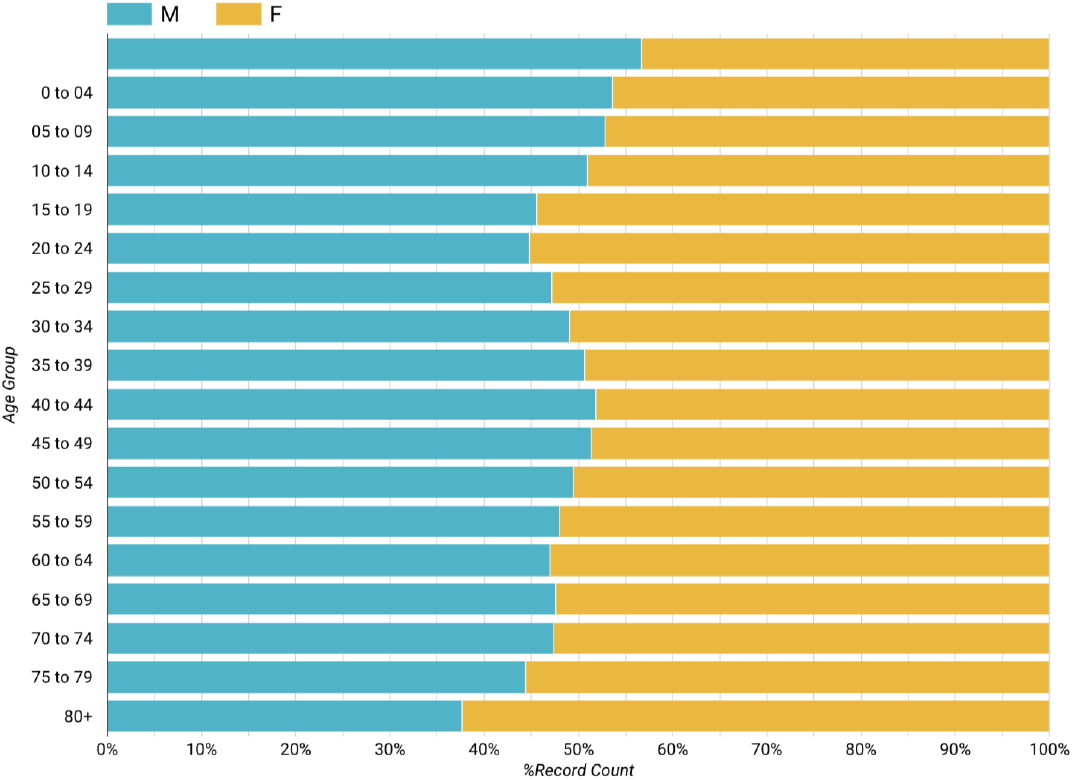
COVID-19 Positive Cases per Sex and Age Group in the Philippines. The x-axis represents the percentage of COVID-19-positive cases, while the y-axis denotes the age group.

Figure 18 provides insight into the distribution of recovered and deceased patients based on sex. Notably, 48.4% of the recovered patients are male (blue), while 51.6% are female (yellow). In contrast, a majority of the patients who succumbed to COVID-19 are male (55.18%), with females accounting for 44.82% of the total fatalities.

**Figure 18:**
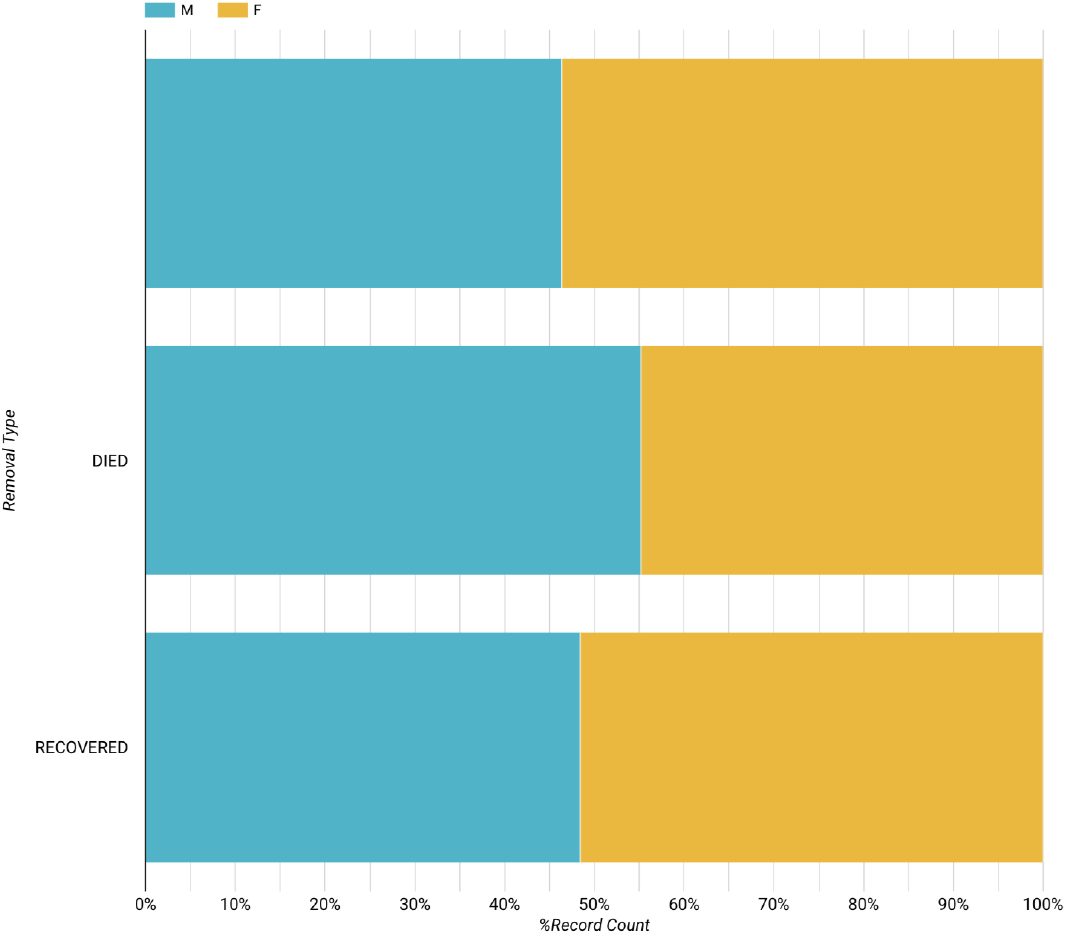
COVID-19 Death Cases and Recovered Cases per Sex in the Philippines. The x-axis represents the percentage of record count of cases, while the y-axis denotes the dead individuals and recovered patients. Male (M) and Female (F) are depicted in blue and yellow, respectively.

Displayed in Figure 19 are the cases of dead individuals (dark gray) and recovered patients (pink) across various age groups. Notably, numerous age groups exhibit a notably high recovery rate. However, a notable number of fatalities are observed in age groups beyond 50, with a significant 12.28% within the 80 and above age group.

**Figure 19:**
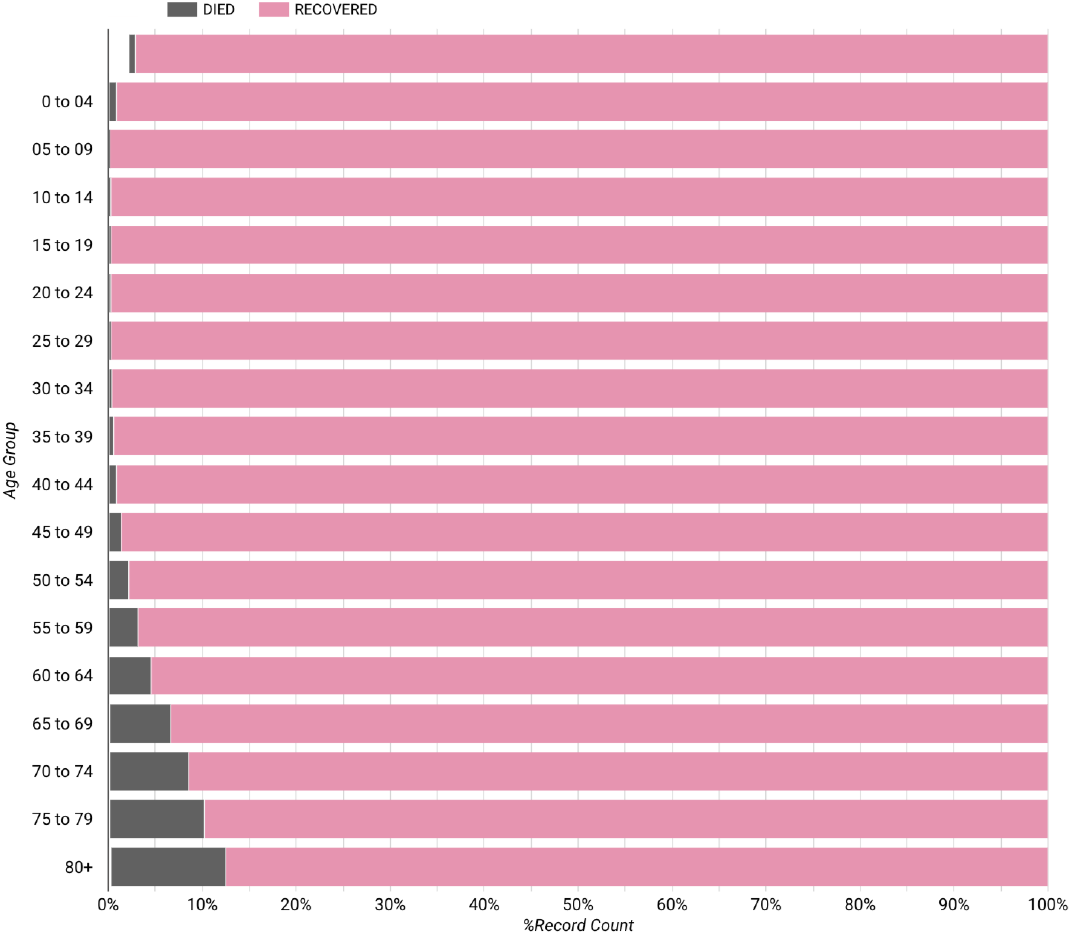
COVID-19 Death Cases and Recovered Cases per Age Group in the Philippines. The x-axis represents the percentage of COVID-19-positive cases, while the y-axis denotes the age group. Died individuals and recovered patients are depicted in dark gray and pink, respectively.

Figure 20 presents the ten provinces, including NCR, with the highest cumulative COVID-19 cases from 2020 to 2023. Notably, the NCR takes the lead with the highest cumulative cases at 1,310,211, followed closely by several provinces in Region 4A, including Cavite (243,409), Laguna (182,174), Rizal (159,121), and Batangas (102,951). In the Visayas region, both Cebu (153,078) and Iloilo (92,336) secure positions within the top 10 provinces. Meanwhile, among the provinces in Mindanao, Davao del Sur (101,478) stands as the sole representative with a significant cumulative case count. This data underscores the varied impact of the pandemic across different provinces in the Philippines, where urban areas tend to have more case counts than rural ones.

**Figure 20:**
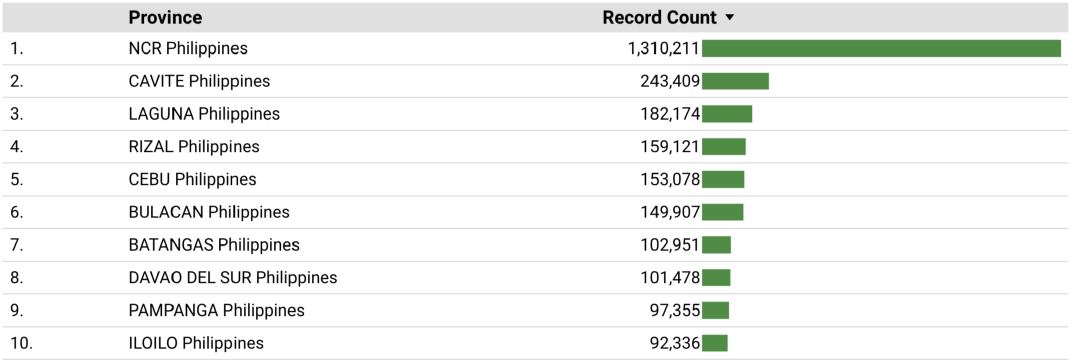
Top Ten Provinces (including NCR) in the Philippines with High COVID-19 Cumulative Cases.

Furthermore, Figure 21 offers a visual representation of the spatial distribution of COVID-19 cases across the country, ranging from areas with high case numbers (red) to those with fewer cases (green). The NCR and Region 4A are prominently marked in red, indicating a high incidence of cases. Region 3 (Central Luzon), Cebu, Iloilo, and Davao del Sur fall within the moderate case range (orange). Region 4A and Region 3 are spatially adjacent to NCR. Conversely, the majority of regions are characterized by low case numbers, denoted by the prevalence of green hues.

**Figure 21:**
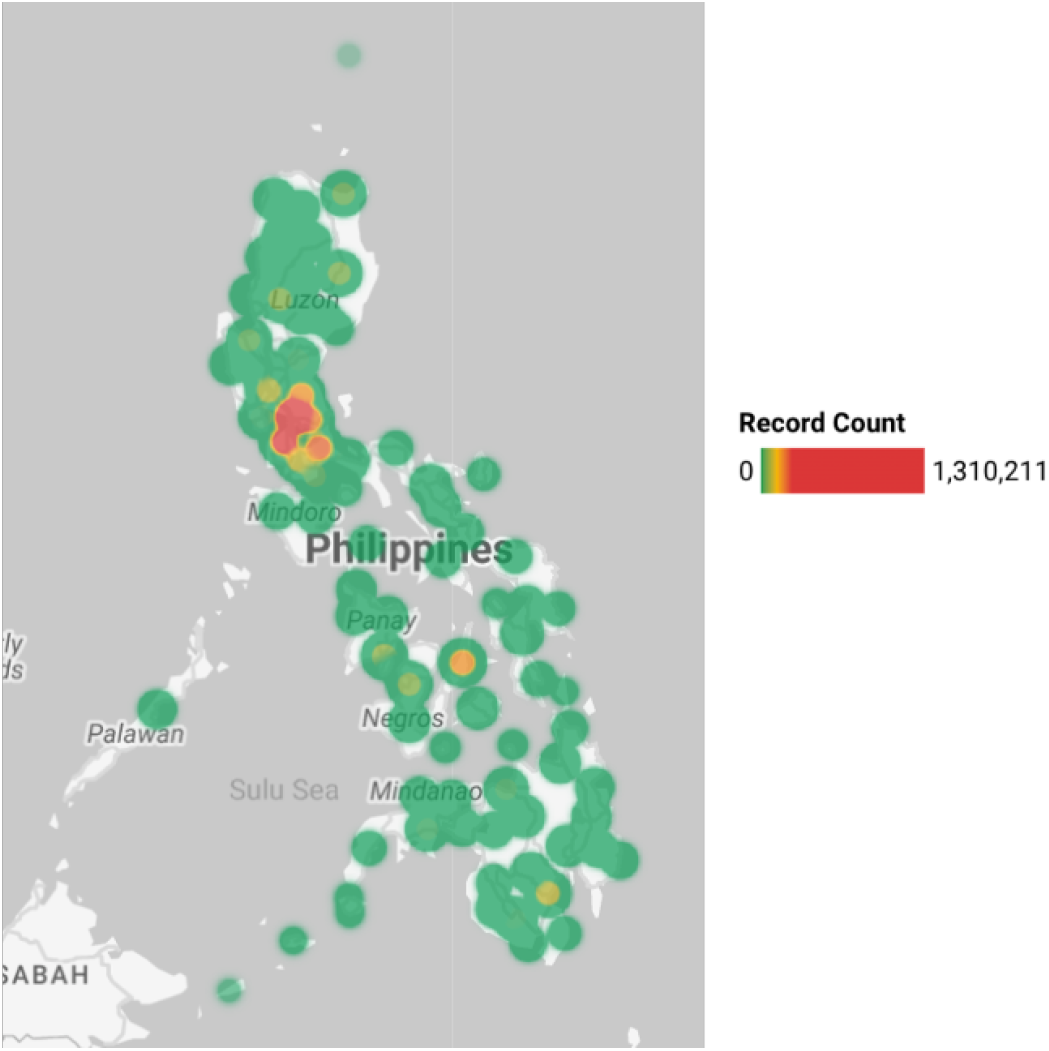
Spatial Distribution of COVID-19 Cases in the Philippines.

Figure 22 depicts the count of individuals who underwent COVID-19 testing (green) alongside those who tested positive for the disease (blue). Notably, the period from April 2020 to September 2021 witnessed a steady rise in the number of individuals tested, reaching an approximate peak of 85,000. Subsequently, from October 2021 to April 2023, the testing figures gradually declined, except for a notable spike in January 2022, where the count surged to around 86,000 individuals. This trend shift could potentially be attributed to the emergence of the Omicron variant of COVID-19. Additionally, in Figure 22, a clear correlation emerges between the number of COVID-19-positive individuals and the count of those tested. Notable peaks in positive cases are observed in September 2021 (24,320 cases) and January 2022 (41,139 cases), aligning with the testing peaks during those periods.

**Figure 22:**
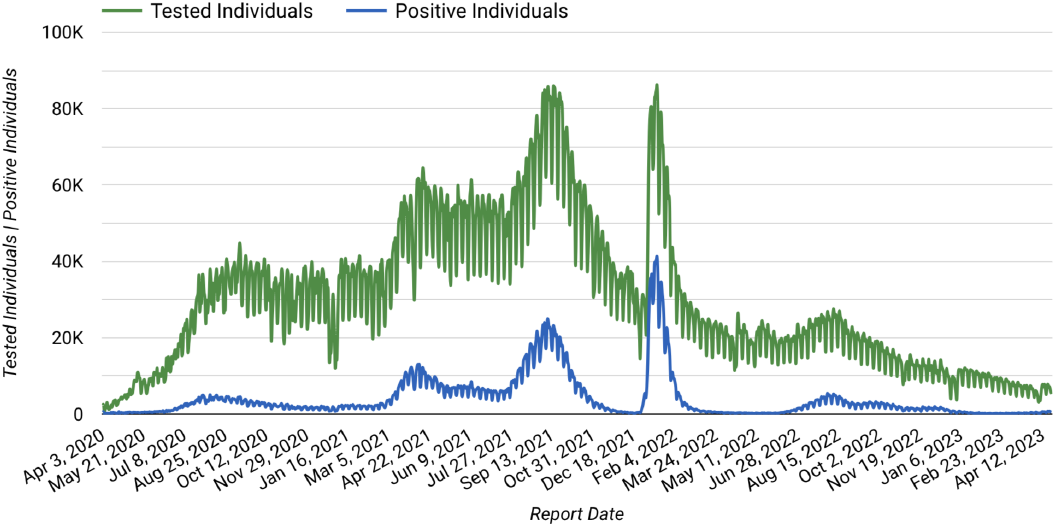
COVID-19 Tested Individuals and Positive Individuals in the Philippines. The x-axis represents the report date of the testing facilities from 2020 to 2023, while the y-axis denotes the number of tested (green) and positive (blue) individuals.

Furthermore, Figure 23 presents the positivity rate among individuals who underwent testing. Notably, instances of a positivity rate exceeding 20% are evident in April, August, and September 2021. Noteworthy is the substantial peak in the positivity rate, reaching 48% in January 2022. A high positivity rate in the early phase of the outbreak could be due to the low number of available testing facilities and test kits.

**Figure 23:**
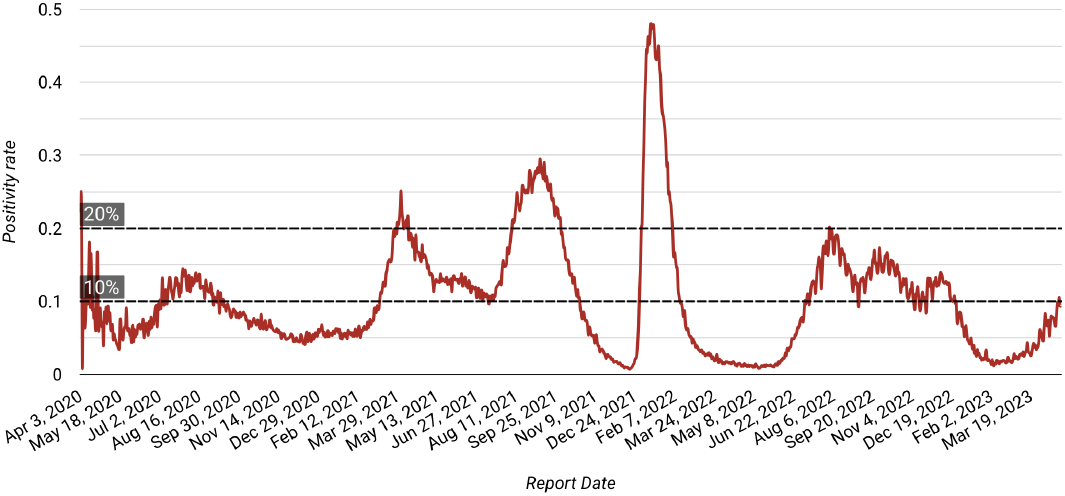
COVID-19 Positivity Rate in the Philippines. The x-axis represents the report date from 2020 to 2023, while the y-axis denotes the COVID-19 positivity rate (red). References are marked at 10% and 20% rates.

Figure 24 illustrates the vaccination landscape within the country. Notably, following the initiation of vaccine distribution, daily vaccinations (depicted in orange) exhibited a significant surge from March 1, 2021, to December 2021. On December 2, 2021, a peak of 1,629,125 individuals received vaccinations. However, post-December 2021, the daily vaccination rate experienced a gradual decline, continuing through February 2023. Moreover, the cumulative count of vaccinated individuals (depicted in blue) demonstrated exponential growth from March 1, 2021, to December 2021, with a total of 56,110,301 people receiving vaccines by December 2, 2021. Similarly, the cumulative number of fully vaccinated individuals (cyan) exhibited exponential expansion from March 1, 2021, to February 21, 2022, reaching a total of 62,652,101 fully vaccinated indi-viduals (more or less 55% of the total Philippine population). The term ‘fully vaccinated’ pertains to those who have completed their primary vaccine series or have received their initial two doses of the COVID-19 vaccine [30]. Additionally, the cumulative count of individuals who have received total booster doses (pink) remains notably lower in comparison to the cumulative fully vaccinated count. As of March 9, 2023, the cumulative figures are as follows: 78,480,056 vaccinated individuals, 74,049,515 fully vaccinated individuals, and 21,756,899 individuals who have received total booster shots.

**Figure 24:**
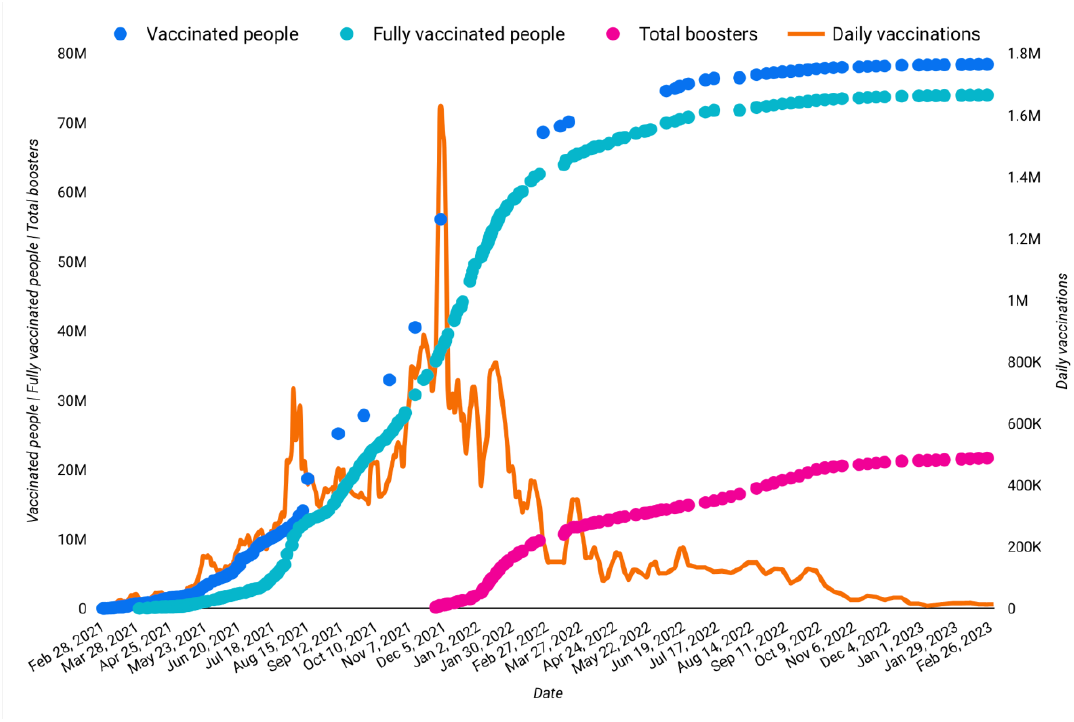
COVID-19 Vaccination Status in the Philippines. The x-axis represents the report date from 2020 to 2023, while the left y-axis denotes the vaccinated people (blue), fully vaccinated people (cyan), and total boosters (pink), and the right y-axis denote the daily vaccinations (orange).

## 3. Philippine COVID-19-related Scientific Papers

With the alarming surge of COVID-19 cases observed in the Philippines, a substantial increase in dedicated research efforts has been witnessed to comprehend this unprecedented pandemic. A multitude of scientific papers have been published, diligently exploring various dimensions of the virus, encompassing its transmission dynamics, impact on public health and education, and potential mitigation strategies.

The government outsourced specific groups, such as the FASSSTER team and the University of the Philippines Pandemic Response team, to devise mathematical model of transmission dynamics and data analytics studies to guide the country’s COVID-19 pandemic response [31]. Researchers have harnessed mathematical models to optimize the allocation of test kits among nationwide testing centers [32]. Mathematical models have adeptly characterized and predicted the transmission dynamics of the disease, offering vital insights into its spread and evolution across diverse populations [33] [34] [35] [36] [37] [38] [39] [40] [41]. Such models have been instrumental in forecasting COVID-19 cases, providing projections essential for policy-makers and healthcare professionals [42] [43] [44].

Beyond transmission and forecasting, mathematical models have played a pivotal role in the vaccination landscape. They have guided the optimal distribution of vaccines [45] [46] [47], assessed the impact of vaccinations on controlling transmission [48], and determined optimal vaccination site selection [49].

In addition to models centered around vaccination, mathematicians have employed their expertise to assess the infection risk faced by frontline healthcare workers [50] and to formulate optimal strategies for controlling the disease [51]. Mathematical models have proven invaluable in quantifying the economic losses stemming from the disease’s propagation and the imposition of lockdown measures within the country [52]. Furthermore, the utility of agent-based models has come to the forefront, enabling the simulation of specific scenarios and the subsequent evaluation of their potential outcomes. Notably, these models have been leveraged to scrutinize the intricate transmission dynamics within classrooms [53], simulate scenarios revolving around the resumption of classes and the implementation of vaccination efforts [54] [55], and provide insights into the impact of the food relief system in mitigating the disease’s effects [56].

In addition to mathematics-driven insights, a myriad of other studies have contributed diverse perspectives to the understanding of the pandemic’s impact. Researchers have delved into the challenges, opportunities, and treatment considerations for diseases such as cancer [57] [58] [59], diabetes [60], HIV [61] [62], and dengue [63] [64]. Further studies have meticulously examined epidemiological and clinical characteristics of COVID-19 patients, furnishing critical insights for public health responses [65] [66] [67] [68]. Some investigation delved into the potential efficacy of utilizing carbon dioxide (CO2) monitors to mitigate the transmission of COVID-19 [69]. A substantial focus has also been directed towards comprehending and addressing the mental health ramifications arising from the pandemic [70] [71] [72].

Education has emerged as a critical focal point, with researchers probing the pandemic’s impact on the education system. Studies have unveiled the multifaceted challenges encountered by students [73] [74] [75] [76], teachers [77] [78] [79] [80] [81], educational institutions [82] [83] [84], and parents [85] [86]. Specific focus has been on the challenges of remote learning [87] [88] [89] and barriers encountered in online learning [90] [91] [92] [93] [94]. Moreover, studies have delved into the cost-benefit aspects of face-to-face school closures [95] and students’ mental well-being [96] [97].

An array of published papers have probed diverse facets of the pandemic’s impact. More studies have explored the psychological influence of the pandemic [98] [99] [100] [101] [102] [103] [104] and its economic repercussions [105] [106] [107]. Research papers presented examinations of government responses to COVID-19 management [108] [109] [110] [111] [112] [113], vaccine hesitancy, and challenges within the vaccination campaign [114] [115] [116] [117] [118] [119] [120].

Figure 25 provides a visual representation of the most frequently recurring words in the titles and abstracts of the 170 selected scientific papers. Among the vast repertoire of 36,055 documented words, ‘COVID’ emerges as the most prevalent term, occurring 643 times. Following closely are the keywords: ‘Philippines’ (379), ‘pandemic’ (239), ‘health’ (228), ‘study’ (159), ‘model’ (115), and ‘students’ (110). These words encapsulate the multifaceted dimensions and concerted efforts of researchers in grappling with the challenges posed by this pandemic.

**Figure 25:**
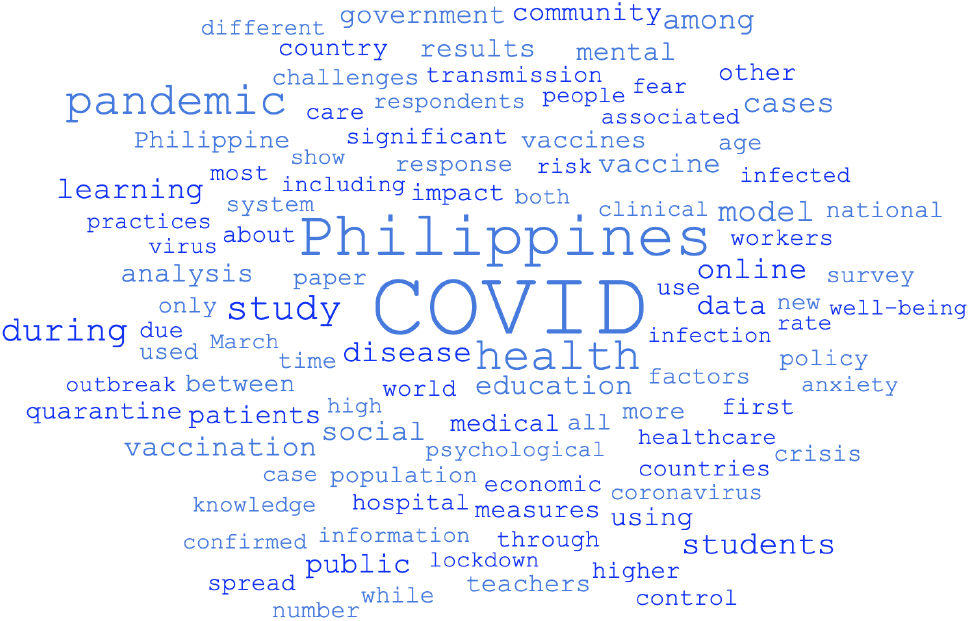
Most Frequent Words in Selected Scientific Papers in the Philippines.

## 4. Remembering the Philippine COVID-19 Time-line

In response to the emergence of COVID-19 in Wuhan City, China, the Philippines’ Inter-Agency Task Force for the Management of Emerging Infectious Diseases (IATF-EID) was promptly convened by the DOH on January 28, 2020 [121]. In alignment with the IATF-EID’s recommendations, the Bureau of Immigration swiftly suspended the issuance of visas upon arrival for Chinese nationals, a measure aimed at curbing group tours from entering the country [122]. On January 29, the Research Institute for Tropical Medicine acquired confirmatory test kits for COVID-19, an essential development due to the absence of local testing capacity. Consequently, the DOH began sending suspected case samples to the Victorian Infectious Disease Reference Laboratory (VIDRL) in Australia [123]. It was on January 30 that VIDRL confirmed the Philippines’ first COVID-19 case, a 38-year-old Chinese woman originating from Wuhan [124]. Swiftly following this revelation, President Rodrigo Roa Duterte (PRRD) enacted a temporary travel ban on individuals from the Chinese province of Hubei [125]. As the virus’s global spread gained momentum, the World Health Organization (WHO) recognized the gravity of the situation by declaring COVID-19 a public health emergency [3] [5].

The first death case of COVID-19 outside China was reported in the Philippines on February 2, 2020 [6]. A month later, in response to the confirmation of the first case of local transmission, the DOH elevated the country’s alert level to Code Red [7]. Acting upon the DOH’s counsel, PRRD formally endorsed a proclamation declaring a state of public health emergency across the nation [126]. Simultaneously, classes at all levels within the NCR were suspended [127]. Concurrently, the Food and Drug Administration (FDA) issued approval for the utilization of the COVID-19 testing kit developed by the University of the Philippines National Institutes of Health [128].

On March 11, 2020, a 67-year-old woman was reported as the first Filipino Citizen in the Philippines to die of COVID-19 [129]. The next day, PRRD announced the expansion of travel restrictions to all countries with local virus transmission [130]. He also imposed a lockdown on Metro Manila - a ban on land, domestic air, and sea travel to and from the region; a prohibition on mass gatherings; and a suspension of classes and flexible work arrangements [16]. With the relentless surge in case numbers, PRRD escalated the response by declaring an enhanced community quarantine (ECQ) for the entirety of Luzon. This tougher quarantine (a.k.a. lockdown) requires strictly observing home quarantine, suspends transportation, and regulates the provision of food and essential healthcare services [17]. Moreover, the gravity of the situation led to the declaration of a state of calamity for the entire Philippines [131]. Furthermore, on April 2, 2020, the authorities mandated all individuals in Luzon to wear face masks or any form of facial protective gear whenever they leave their homes (e.g., face shields) [132].

In direct response to the lockdown measures, the University of the Philippines (UP), the nation’s premier university, exhibited the core tenets of ‘Bayanihan’. United by this spirit, UP and its constituent communities swiftly rallied to assist stranded students. Notably, ‘Oplan Kawingan’ was instituted at UP Los Baños (UPLB), while the UP Diliman University Student Council introduced ‘IskoOps: COVID-19.’ Additionally, the ‘UPLB’s Oplan Hatid’ initiative was initiated, ensuring the secure transportation of UP students to their respective family homes [133].

The UP COVID-19 Pandemic Response Team (UP PRT), primarily composed of educators, researchers, statisticians, mathematicians, and social scientists across various UP campuses, has conducted simulations to forecast the peak of COVID-19 cases within Metro Manila. Drawing upon their mathematical models, this expert team projected that the peak of cases (first wave) was anticipated between late April and June 2020, encompassing a projected range of 140,000 to 550,000 infected individuals in Metro Manila. UP PRT strongly emphasized the importance of community collaboration in the collective effort to combat the virus and effectively flatten the epidemic curve [134] [135] [136]. Dr. Jomar Rabajante, a UPLB mathematics professor and UP PRT fellow, developed an SEIR model tailored for Metro Manila. His projections indicated a peak in mid-April, with around 2,500 active COVID-19 cases daily. Dr. Rabajante accentuated the significance of long-term planning, cautioning against the potential resurgence of infections should regular activities resume following the ECQ. He proffered the notion of transitioning from national lockdown to a localized modified community quarantine with well-defined government strategies implemented at the LGU level [137] [134] [136].

The time-series analysis of UP PRT showed a longer doubling time for confirmed cases and reported a reproduction number below 1, indicating the ECQ’s effectiveness. As a result, they proposed a graduated activation of the ECQ after April 30, 2020, based on the level of risk in specific areas [138] [139]. In addition, UP PRT models also indicated that opening all schools in NCR may increase COVID-19 transmission. UP PRT’s analysis indicated that individuals aged 0-20 engage in the highest levels of social interaction. These findings supported the government’s cautious approach to school reopening [140] [141]. Nevertheless, a study conducted by the Asian Development Bank, with Dr. Rabajante as co-author, also highlighted the costs due to school closures [95].

On May 12, 2020, the UP PRT raised concerns about errors and inconsistencies in the DOH’s COVID-19 patient data, emphasizing the need for accurate and timely information [142]. This event initiated discussions and the eventual improvement in COVID-19 data reporting.

Two months after the strict lockdown, the government has decided to gradually ease the quarantine restrictions in certain areas. The regions of Metro Manila, Laguna province, and Cebu City, which were classified as “high risk” communities by the IATF-EID, were placed under a modified enhanced community quarantine (MECQ), while the remaining areas in the country were under a general community quarantine (GCQ). Under the MECQ, some businesses were allowed to partially operate, and limited outdoor exercises were permitted. However, public transportation was still prohibited. On the other hand, during the GCQ, public transportation was allowed to operate but with reduced capacity [143] [144] [145].

PRRD transitioned NCR quarantine from MECQ to GCQ despite COVID-19 challenges. Effective June 1, 2020, due to the slowing infection rate and the mayors’ request, more nonessential businesses reopened to revitalize the economy and support workers [146]. However, in contrast to this development, Cebu City has reverted to the strictest form of ECQ due to a surge in COVID-19 cases [147]. The UP PRT cautioned against prematurely easing quarantine measures in NCR based on data from the DOH. Projections from the UPLB Biomathematics Team suggested that the country’s coronavirus cases could reach as high as 95,000 by the end of August 2020 [148]. Furthermore, the UPLB Biomathematics Team has developed two free modeling tools: the Job Risk Calculator and the Workplace Outbreak Simulator [149]. At the same time, the UP PRT has launched a web portal named endcov.ph which includes features like case projections, density maps of active cases, and CITY vs. COVID [150] [151].

As of July 13, 2020, five officials in the Philippines were leading the fight against the coronavirus as anti-COVID czars [152]. The testing czar, Vince Dizon, introduced Oplan Kalinga for patient transport and announced that a negative swab test was no longer required for declaring COVID-19 patients as recovered [153]. On the other hand, PRRD announced that an MECQ would be imposed in Metro Manila and four nearby provinces from August 4 to 18, 2020 [154]. At the same time, on August 4, 2020, former PhilHealth official Thorrsson Montes Keith accused top officials of embezzling Php 15 billion through fraudulent schemes. PhilHealth CEO Ricardo Morales denied the allegations and questioned Keith’s credibility [155]. Furthermore, the Philippine economy plummeted in Quarter 2 as COVID-19 lockdowns hit hard [156].

As of August 6, 2020, as per DOH, there were 100 licensed COVID-19 testing laboratories in the Philippines [157]. On August 16, 2020, the G614 variant of SARS-CoV-2 was detected in the Philippines [158].

The law called Bayanihan to Recover as One Act was signed by PRRD on September 11, 2020, allocating a fund of Php 165.5 billion for pandemic response and recovery [159]. On October 5, 2020, over 22 million students from public schools returned to school in the Philippines without face-to-face classes, according to Education Secretary Leonor Briones [160]. Meanwhile, Albay 2nd District Rep. Joey Salceda announced on October 16, 2020, that the proposed Php 4.5-trillion national budget for 2021 in the Philippines includes Php 893 billion for the pandemic response [161].

On November 19, 2020, PRRD agreed to advance supply agreements and make early payments to COVID-19 vaccine manufacturers [162]. Moreover, the government has mandated (but not well-received by the public) the use of the StaySafe contact tracing app to strengthen the COVID-19 response in high-infection areas [163]. Furthermore, before the end of 2020, the Philippines suspended flights from the UK due to a new coronavirus strain [164].

Several LGUs, as of January 11, 2021, had successfully secured vaccine doses for their constituents [165] [166]. On January 13, 2021, the DOH reported that the Alpha variant of the coronavirus, believed to be more contagious, has been detected in the Philippines [167].

On January 25, 2021, the Commission on Higher Education approved limited face-to-face classes for medicine and allied health sciences students in areas under MGCQ [22]. Moreover, the government allowed the reopening of cinemas, game arcades, and leisure activities in GCQ areas on February 12, 2021 [23]. In late February, the government received its first shipment of COVID-19 vaccines from Sinovac Biotech [8].

The Philippines started its COVID-19 vaccination drive on March 1, 2021, with the first doses of Sinovac’s CoronaVac given to the Philippine General Hospital Director and a nurse. Other officials also received the vaccine [168] [169]. On the 2nd day of March, the Philippines detected its first six cases of the Beta variant of COVID-19 [21]. Furthermore, the UP Philippine Genome Center, National Institutes of Health, and DOH conducted biosurveillance and identified a concerning SARS-CoV-2 variant in the Philippines named Lineage P.3. The detection was based on 33 samples collected mainly from Central Visayas [170] [171].

Starting March 28, 2021, the Philippine government vaccinated senior citizens and people with comorbidities alongside healthcare workers [172]. On the other hand, due to the significant rise in COVID-19 cases, starting March 29, stricter measures were implemented in the Greater Manila area (ECQ) until April 4 [173].

On May 11, 2021, the Philippines confirmed its first two cases of the COVID-19 Delta variant [174]. Moreover, the country confirmed local transmission of the Delta variant of COVID-19 on July 22, 2021 [25]. Due to the surge of the Delta variant, PRRD has approved the recommendation to place Metro Manila, Laguna, and Bataan under MECQ until the end of August [26]. Dr. Rabajante said that COVID-19 cases in the Philippines were spreading rapidly, with daily infections potentially reaching 19,000 if the current trend continues [175].

After a month of strict lockdown, Metro Manila shifted to GCQ and serve as a pilot area for granular lockdowns from September 8 to the end of the month. Starting September 16, Metro Manila was under Alert Level 4, with granular lockdowns enforced in critical areas. The alert level system focuses on targeted restrictions and allows economic activities beyond the lockdown areas [176] [177].

On September 6, the Philippine government launched VaxCertPH, a digital COVID-19 vaccination certificate system developed by the Department of Information and Communications Technology (DICT) [178]. PRRD has approved the vaccination of the general population, including minors, against COVID-19 starting in October 2021 [27]. The pandemic situation in the country was then improving, indicated by declining daily cases, hospitalizations, and positivity rate, according to Dr. Rabajante. However, the reported numbers may not reflect the actual daily infection due to possible underreporting, as antigen test results were not included in the official case tally [179].

On October 2021, Go Negosyo and OCTA Research partnered to analyze COVID-19 data, guiding micro, small, and medium enterprises (MSMEs) with insights for safe economic reopening. OCTA becomes Go Negosyo’s COVID-19 advisory, crafting data-driven strategies and sharing pandemic response lessons. They influenced Metro Manila’s Alert Level 3 downgrade and propose scientific alert level methods based on hospitalization rates and metrics [180].

The COVID-19 Alert Level System (ALS) was implemented starting October 20, replacing the imposed quarantine classifications. Some areas were placed under specific alert levels based on their COVID-19 situation [181]. On November 11, 2021, PRRD approved the nationwide implementation of an ALS, and on November 22, the ALS was fully implemented nationwide [182] [183]. On the other hand, the Philippine government launched a COVID-19 booster campaign for seniors and immunocompromised individuals, authorizing additional doses for both groups [184].

The Philippines confirmed its first two cases of the Omicron variant in December 2021 [28]. Dr. Rabajante predicted that with the emergence of the Omicron variant and increased mobility due to the holidays, the daily COVID-19 cases in the country could escalate to 40,000 by mid-January [185]. On January 6, 2022, the Department of the Interior and Local Government (DILG) urged the public not to be involved in panic buying amid reported shortages of medicines like paracetamol [186].

On February 1, 2022, fully vaccinated Filipinos and foreigners from visa-free countries can enter the Philippines for tourism or business purposes. Mandatory quarantine was no longer required, only a negative RT-PCR test within 48 hours before departure [187]. Moreover, from January to May 2022 experienced huge mass rallies in support of candidates running for positions in the May 2022 national elections. There was no significant increase in COVID-19 cases during the election season.

As the Philippines entered its third year of battling the pandemic, the government began vaccinating children aged 5 to 11 against COVID-19 on February 7 [27]. After a month, the Philippines shifted from providing daily COVID-19 updates to weekly updates as the country entered the “new normal” under Alert Level 1 [188]. On May 18, 2022, senior citizens and frontline health workers in the Philippines can already receive a second booster shot of mRNA vaccines [9]. As of June 19, 2022, over 70 million people in the Philippines have been fully vaccinated against COVID-19 [189].

On July 5, 2022, minors aged 12 to 17 in the Philippines were now eligible for COVID-19 booster shots, according to the DOH [190]. To increase the vaccination rate, the DOH has launched the “PinasLakas” campaign, a nationwide booster vaccination drive in the Philippines [191].

From being required, the Cebu City government implemented a voluntary face mask policy on September 5, 2022 [192]. After one month, President Ferdinand Marcos Jr. implemented a policy where wearing face masks indoors and outdoors became optional in the country. However, wearing face masks was still advised for specific groups and areas [193]. On the other hand, on October 18, 2022, the DOH announced the detection of the Omicron XBB subvariant and XBC variant of COVID-19 in the country [194].

In November 2022, Malacañang approved the IATF-EID’s recommendation to ease testing and quarantine pro-tocols for inbound travelers to the Philippines amid the COVID-19 pandemic [195]. On March 7, 2023, the Department of Tourism (DOT) announced that the wearing of face masks and proof of full COVID-19 vaccination was no longer required in tourist spots in the Philippines [196]. Furthermore, DOH announced that there was no need to fear the recent 19% increase in new COVID-19 cases [197] as the percent increase was also coming from a very low base count, and the number of severe cases is still far from risky levels.

On April 12, 2023, the DOH announced that the general population in the Philippines was now eligible to receive the second COVID-19 booster shots [198]. Eleven (11) vaccines have been approved for use (under Emergency Use Authorization) in the Philippines, including COVOVAX, Comirnaty, Spikevax, Sputnik Light, Sputnik V, Jcovden, Vaxzevria, Covaxin, Covilo, Inactivated (Vero Cells), and CoronaVac [199].

On May 5, 2023, the WHO Director-General transmitted the report of the 15th meeting of the IHR Emergency Committee on the COVID-19 pandemic. The Committee highlighted decreasing COVID-19 deaths, hospitalizations, ICU admissions, and high population immunity. The Director-General agreed with the Committee’s advice and declared that COVID-19 was no longer a public health emergency of international concern [200]. In July 2023, President Ferdinand Marcos Jr., with the support of DOH, lifted the nationwide state of public health emergency due to COVID-19 [201] [202].

See the Supplementary Information for more references of selected events that are important to be noted as part of the Philippine COVID-19 timeline.

## 5. Conclusion

This study presents an in-depth descriptive analysis of COVID-19 data in the Philippines, a middle-income country, spanning the period from 2020 to 2023. The findings highlight the notable concentration of cumulative COVID-19 cases in the NCR and CALABARZON. The observed surges in cases were primarily attributed to the emergence of COVID-19 variants and the gradual easing of quarantine protocols and measures. As of April 21, 2023, the country has reported a total of 4,087,964 COVID-19 cases and 66,444 related deaths [11]. Notably, on July 22, 2023, following the termination of the public health emergency declaration by President Ferdinand Marcos Jr., the DOH lifted all medical protocols on COVID-19 in the country [201] [202].

The core objective of this paper is to retrospectively enumerate the highlights of the COVID-19 pandemic in the Philippines, facilitating a forward-looking perspective and aiding the nation’s journey in the ‘new normal.’ By curating a selection of pertinent scientific papers and creating a chronological timeline of significant events during the pandemic as well as visualizing the COVID-19 datasets, we aim to provide a comprehensive understanding of the multifaceted challenges and responses.

Moreover, this study underscores the pivotal role played by mathematicians, statisticians, and data scientists, who have emerged as key contributors amid the crisis. Their mathematical models have yielded crucial insights into various facets of the pandemic, from transmission dynamics to vaccination strategies and impact assessment.

This paper stands as a potential cornerstone reference, offering a concise but comprehensive summary of the Philippines’ battle against COVID-19. Furthermore, it serves as a valuable resource for gaining insights into this unprecedented global challenge, guiding the decision-making processes of policymakers and public officials in navigating the uncharted waters of the future.

## Data Availability

All data produced are available online at World Health Organization (https://covid19.who.int/);
Department of Health of the Philippines (https://doh.gov.ph/covid19tracker);
Our World in Data (https://ourworldindata.org/covid-vaccinations);
Google, Covid-19 community mobility reports, (https://www.google.com/covid19/mobility/)

## 6. Supplemental Information

Some references for the timeline of selected events during the COVID-19 pandemic can be found in the Supplementary Information.

